# Identification of novel nutrient sensitive human yolk sac functions required for embryogenesis

**DOI:** 10.1101/2024.09.30.24314635

**Authors:** M White, J Arif-Pardy, E Bloise, KL Connor

## Abstract

The human yolk sac (hYS) is essential for embryo nutrient biosynthesis/transport and development. However, there lacks a comprehensive study of hYS nutrient-gene interactions. Here we performed a secondary analysis of hYS transcript profiles (n=9 samples) to identify nutrient-sensitive hYS genes and regulatory networks, including those that associate with adverse perinatal phenotypes with embryonic origins. Overall, 14.8% highly expressed hYS genes are nutrient-sensitive; the most common nutrient cofactors for hYS genes are metals and B-vitamins. Functional analysis of highly expressed hYS genes reveals that nutrient-sensitive hYS genes are more likely to be involved in metabolic functions than hYS genes that are not nutrient-sensitive. Through nutrient-sensitive gene network analysis, we find that four nutrient-sensitive transcription regulators in the hYS (with zinc and/or magnesium cofactors) are predicted to collectively regulate 30.9% of highly expressed hYS genes. Lastly, we identify 117 nutrient-sensitive hYS genes that associate with an adverse perinatal outcome with embryonic origins. Among these, the greatest number of nutrient-sensitive hYS genes are linked to congenital heart defects (n=54 genes), followed by microcephaly (n=37). Collectively, our study characterises nutrient-sensitive hYS functions and improves understanding of the ways in which nutrient-gene interactions in the hYS may influence both typical and pathological development.

## Introduction

The human yolk sac (hYS) is a transitory extraembryonic membrane that plays a singular role in orchestrating early embryogenesis [1]. Shorter-lived than other extraembryonic membranes like the amnion and placenta, the hYS originates from the hypoblast layer of the bilaminar embryonic disc around embryonic days (ED) 9-10, continues to grow progressively to form the secondary YS, is functional up to the 10th week of pregnancy, and is no longer identifiable by the 20th week [1]. The hYS contains inner endodermal cells facing the vitelline cavity, mesodermal cells (containing vascular precursors), and an outer mesothelial layer facing the coelomic cavity [2-4]. One of its primary functions is the mediation of embryonic nutrition and protection through transporter-mediated nutrient exchange processes [1, 5, 6]. The hYS forms a biological barrier between the embryonic primitive gut and the coelomic fluid (CF), the first fluid within the gestational sac [1, 7]. CF contains proteins produced by the villous trophoblast, embryonic tissues, and the secondary YS, and functional analysis of CF proteins suggests a role in nervous system and respiratory processes [8-10]. Based on their anatomical proximity, it is expected that gene networks in the hYS interact directly and indirectly with the CF to regulate embryonic development. A recent global transcriptomic approach to identify hYS genes supports this, finding proteins in the CF that are transcribed for in the hYS [1, 11]. Although hYS transcriptomics and CF proteomics can aid in understanding the transport and biosynthetic functions of the hYS, nutrient-gene interactions within the hYS and the potential implications for embryonic development remain unclear and rarely explored.

It is well-recognised that multiple micronutrients are required to support or mediate early embryogenic processes [12-22]. For example, hYS-related and nutrient-sensitive disease mechanisms for spontaneous abortion, have been suggested, as both abnormal hYS shape and excess copper and/or vitamin A have been associated with spontaneous abortion [23, 24]. Low levels of certain micronutrients, including vitamins B9 (folate) [25, 26], B12 [25], and D [27, 28], zinc [29, 30], methionine [31], iron [32], and magnesium [21], as well as high levels of other micronutrients, like copper and vitamin A [30, 33, 34], have also been associated with the formation of congenital anomalies. However, the mechanisms underlying relationships between nutrient insufficiency or excess and increased risk of congenital anomalies, such as neural tube defects (NTDs), renal agenesis, gastroschisis, orofacial clefts, and congenital heart defects (CHDs), are largely unknown. This is in part due to the ethical difficulty in obtaining hYS tissue for research. As the hYS plays a role in primitive gut formation, neural tube closure, and heart looping, which all occur between 3-7 weeks post-fertilization [1, 35, 36], [37], it is likely that the hYS plays a role in mediating nutrient-sensitive disease risk for the developing embryo. Investigating the global landscape of micronutrient-gene interactions in hYS tissue may provide a better understanding of hYS physiology and improve understanding of the pathogenesis of nutrient-sensitive pathologies that originate during embryogenesis.

The overall aim of this study was to characterise nutrient-gene interactions within the hYS and CF, and to understand the extent to which nutrient-sensitive hYS genes overlap with genes known to associate with perinatal pathologies with embryonic origins. Using a nutrient-focused transcriptomic and proteomic systems approach, we first characterised nutrient-sensitive genes and regulatory networks of the hYS [38]. Next, we described which nutrient transporters were most highly expressed by the hYS, and determined if, and which, nutrient-sensitive hYS genes are expressed as proteins in the CF. Last, we identified nutrient-sensitive genes expressed in the hYS that associate with perinatal pathologies that originate in early development. Collectively, these results provide a global view of micronutrient-gene interactions in hYS tissue and improve our understanding of hYS physiology and the pathogenesis of nutrient-sensitive pathologies that originate during embryogenesis.

## Methods

### Data source and pre-processing

First-trimester hYS RNA sequencing (RNA-seq) data were obtained from a publicly available dataset (n=9 hYS samples) [11]. To characterise a subset of highly expressed hYS genes, we identified genes with ≥10 reads per kilobase of transcript, per million mapped reads (RPKM), as has been used as a cut off in other secondary analyses of RNA-seq data [39-43]. Genes with ≥10 RPKM have been described as abundantly or actively expressed [39, 43], or as expressed highly enough to differentiate cell lines [41]. Three expression groups were defined for subsequent analysis: highly expressed hYS genes (≥10 RPKM), lowly expressed hYS genes (<10 RPKM), and all genes expressed in the hYS.

### hYS nutrient-gene interactions

#### Identifying nutrient-sensitive hYS genes

We applied a nutrient-focused transcriptomic analysis approach that we and other have previously published [38, 44, 45] to identify and describe genes expressed in the hYS whose function is sensitive to the bioavailability of nutrients. A comprehensive list of proteins with nutrient cofactors (n=2301) was obtained from a publicly available dataset and used to identify highly expressed hYS genes encoding for a protein with a nutrient cofactor [45] (hereafter referred to as nutrient-sensitive genes). This dataset was constructed using the EBI CoFactor [46], the Uniprot [47], Expasy [48] and the Metal MACiE [49] databases. The proportion of nutrient-sensitive genes in each of the three expression groups, as well as the nutrient cofactors associated with each gene, were identified.

#### Functional categorization of nutrient-sensitive hYS genes

Functional classification of the highly expressed hYS genes was performed using Protein ANalysis THrough Evolutionary Relationships (PANTHER) software [50]. The Gene Ontology (GO) Molecular Functions and Biological Processes, and PANTHER Protein Classes of highly expressed nutrient-sensitive and non-nutrient-sensitive hYS genes were identified. PANTHER Pathway classifications were also used to identify key gene pathways involving highly expressed nutrient-sensitive hYS genes.

#### Network analysis of nutrient-sensitive hYS gene regulators

To characterise nutrient-sensitive gene networks in the hYS, we next aimed to identify nutrient-sensitive miRNAs and transcription regulators (TRs) that were expressed in the hYS and are known to regulate other highly expressed hYS genes [38].

We first identified hYS genes that encoded a miRNA. Then, using miRwalk (version 2.0) [51], a database that contains information on predicted and validated miRNA-gene (target) interactions, we obtained the targetome files for each highly expressed miRNA in the hYS. Targetome files were filtered to contain only validated miRNA-gene interactions and reviewed against the list of highly expressed genes in the hYS. Last, we identified highly expressed, nutrient-sensitive hYS genes that were targeted by highly expressed hYS miRNAs.

We used iRegulon (version 1.3) to identify possible nutrient-sensitive TRs that regulate highly expressed hYS genes. Through a reverse-engineering approach and cis-regulatory sequence analysis, iRegulon predicts the transcriptional regulatory network of gene lists and may outperform alternative motif-discovery methods [52]. A list of candidate TRs was produced and screened to identify nutrient-sensitive TRs, using the same approach described above to identify nutrient-sensitive hYS genes [45]. Network files of nutrient-sensitive TRs and their target genes in the hYS were exported and used to generate regulatory network diagrams in Cytoscape (version 3.9.1).

### Identifying highly expressed nutrient transporters in the hYS

To identify nutrient transporters expressed by the hYS, we cross-referenced a previously published list of genes encoding nutrient, or nutrient precursor, transporter proteins (n=312 genes) [53] against the list of highly expressed hYS genes. The overlap between the two lists was analysed to identify which nutrient’s transporters are most abundant in the hYS, and which hYS nutrient transporters are nutrient sensitive.

### Nutrient-sensitive proteomic analysis of coelomic fluid

We next determined whether, and which, nutrient-sensitive hYS genes are expressed as proteins in the CF. We collated publicly available human CF proteomic data from two previous studies (n=9 samples [the same study as the hYS transcriptome data]; 165 proteins [11], and n=22 samples; 96 proteins [8]) to generate a list of proteins expressed in the CF (n=248 proteins total, after manually excluding duplicate proteins that appeared in both datasets). CF samples were collected via ultrasound-guided transvaginal puncture from uncomplicated pregnancies at 7- to 12-weeks’ gestation from patients undergoing elective surgical pregnancy termination [11], or between 7- to 10-weeks’ gestation from pregnancies with chromosomally normal fetuses [8].

Highly expressed nutrient-sensitive hYS genes that encoded any of the 248 CF proteins were identified, and pathway analysis was performed using PANTHER to understand the function of hYS genes also expressed in the CF as proteins [50].

### Associations between nutrient-sensitive hYS genes and perinatal pathologies

Lastly, to identify and describe nutrient-sensitive hYS genes that are associated with perinatal pathologies originating during embryogenesis, we first generated a reference list of perinatal pathologies originating during embryogenesis and up to 20 weeks’ gestation (when the hYS completely degenerates [1]). As a starting point, a list of major congenital diseases was obtained from the Centre for Disease Control (CDC) [54]. A literature review was conducted using PubMed to determine the etiological timing of each listed perinatal pathology, and those with a confirmed or plausible etiological origin prior to 20 weeks’ gestation [55-69] were carried forward for further review. Two additional outcomes, “spontaneous abortion” (loss of pregnancy before 20 weeks’ gestation) and “abnormal yolk sac morphology”, were added to the list, as both are associated with adverse perinatal outcomes and originate during the embryogenic period [70, 71]. The reference list of congenital anomalies with confirmed or plausible etiological origin prior to 20 weeks’ gestation included: exomphalos, gastroschisis, NTDs, CHDs, anorectal atresia, renal agenesis, esophageal atresia, orofacial cleft, large intestinal atresia, spontaneous abortion, and abnormal yolk sac morphology (Supplementary Figure S1).

Next, nutrient-sensitive hYS genes associated with the pre-defined congenital anomalies were identified. First, a list of verified gene-phenotype associations, supported by published literature, was generated for each congenital anomaly by collating data from the Comparative toxicogenic database (CTDbase) [72], the Mouse Genome Database (MGI) [73], and the Genome Wide Association Study (GWAS) catalog [74]. Evidence from mouse models on gene associations with congenital anomalies was included here, as much of the foundational research on molecular and cellular contributions to developmental abnormalities is performed in mice. Although translating findings from animal models to humans has limitations, mice are widely used in developmental biology research, due to their mammalian biology and the high conservation of genes, proteins, and regulatory pathways between these species [75], and because mouse and human YS share similar morphologies and functions, including nutrient exchange and metabolism [76]. Each gene list contained gene symbols and the species in which the gene-phenotype association was identified. The list of genes known to associate with each congenital anomaly was then cross-referenced with the list of highly expressed nutrient-sensitive hYS genes, to identify nutrient-sensitive hYS genes that associate with at least one perinatal pathology that originates during embryogenesis.

## Results

### Genes with B vitamin- and metal-cofactors are abundant and highly expressed in hYS

Of the 3648 highly expressed genes in hYS (Supplementary Figure S2), 14.8% (n=540 genes) were nutrient sensitive (Figures 1-2 and Supplementary Table S1). Similarly, 13.2% (n=1164) and 13.7% (n=1704) of lowly expressed hYS genes and all hYS genes, respectively, were nutrient sensitive (Figures 1B-C). The proportion of genes sensitive to specific nutrients was determined; most hYS genes were sensitive to metals and B vitamins, regardless of level of expression (Figure 1 and Supplementary Figure S3).

**Figure 1.**
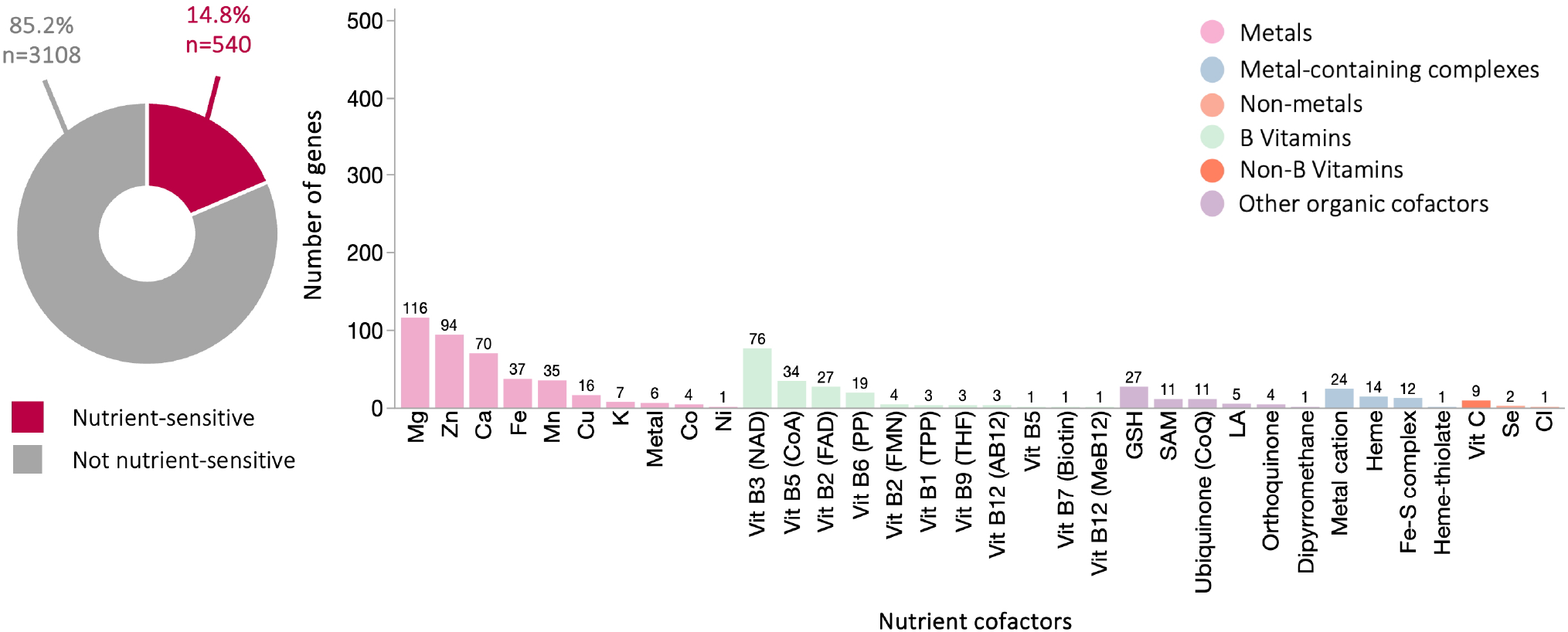
Highly expressed nutrient-sensitive genes in hYS genes (n=3648). Proportion of hYS genes that are nutrient-sensitive (left column; donut charts) to the nutrients indicated in the histograms (right column; bar charts). Numbers above the histogram bars = total number of genes sensitive to that nutrient. RPKM = Reads per kilobase of transcript, per million mapped reads; SAM = S-Adenosyl methionine; GSH = glutathione; Mo = molybdenum; MPT = molybdopterins.

The top 10 most highly expressed nutrient-sensitive hYS genes were collectively sensitive to S-Adenosyl methionine, vitamin B3, iron/heme, copper, zinc, calcium, and potassium (Figure 2). Six of the top 10 most highly expressed nutrient-sensitive hYS genes were mitochondrially encoded: Mitochondrially Encoded NADH Dehydrogenase 4L (MT-ND4L), Mitochondrially Encoded NADH 3 and 4 (MT-ND3 and MT-ND4), Mitochondrially Encoded Cytochrome B (MT-CYB), and Mitochondrially Encoded Cytochrome C Oxidase I and II (MT-CO1 and MT-CO2).

**Figure 2.**
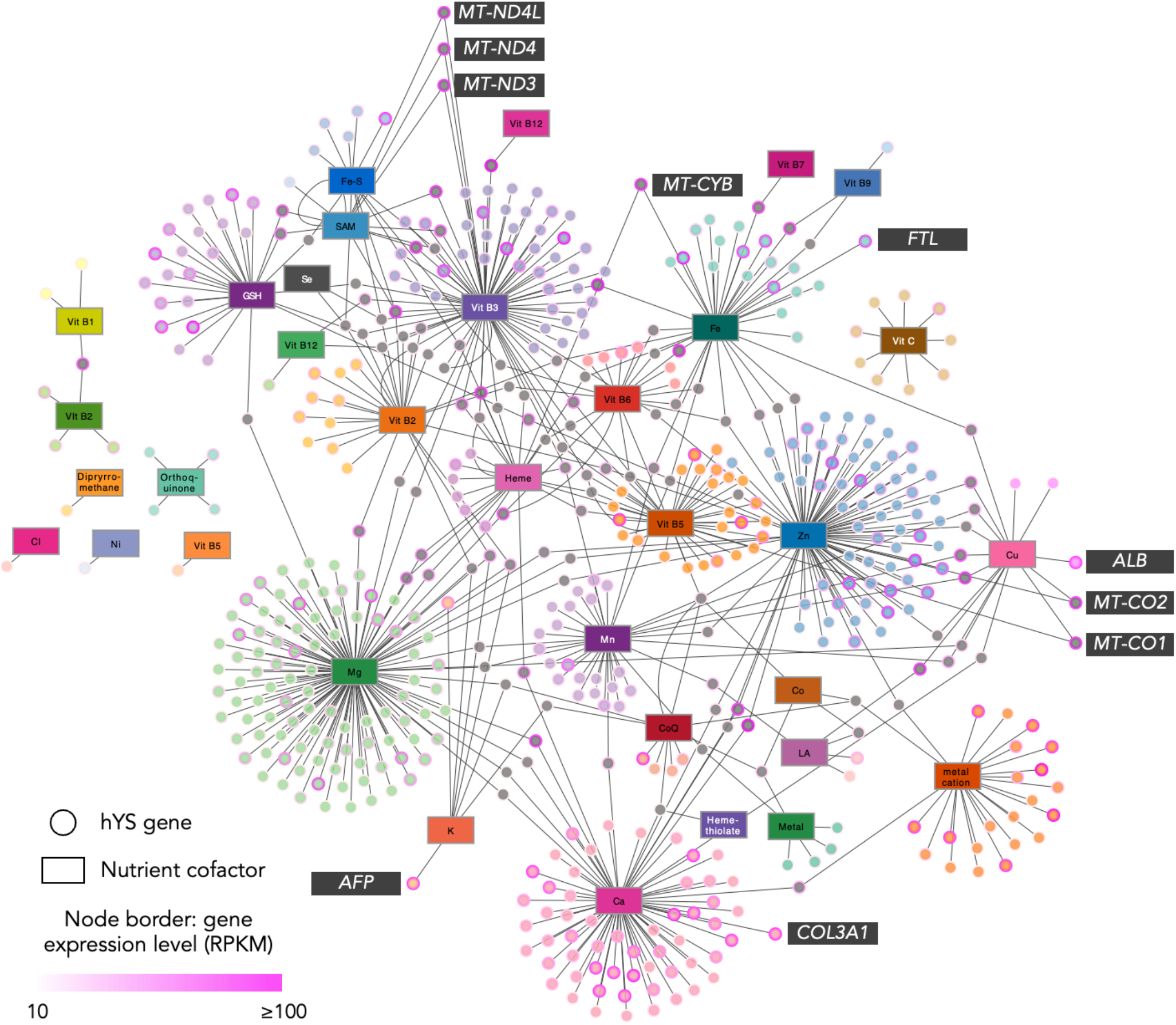
Nutrient-gene interactions in hYS. Nutrient-sensitive genes (circle nodes) connected to their nutrient cofactors (rectangular nodes) via edges (lines). Genes with one nutrient cofactor are colour-coordinated to their respective cofactor. Genes (nodes) with >1 nutrient cofactor are grey. The top 10 most highly expressed hYS are labelled. *MT-ND4L* = Mitochondrially Encoded NADH Dehydrogenase 4L; *MT-ND3/4* = Mitochondrially Encoded NADH; *FTL* = Ferritin Light Chain; *MT-CYB* = Mitochondrially Encoded Cytochrome B; *MT-CO1* = Mitochondrially Encoded Cytochrome C Oxidase; *ALB* = Albumin; *MT-CO2* = Mitochondrially Encoded Cytochrome C Oxidase II; *COL3A1 =* Collagen Type III Alpha 1 Chain; *AFP* = Alpha Fetoprotein; RPKM = reads per kilobase of transcript, per million mapped reads. Vit = Vitamin; CoQ = ubiquinone.

### Functions of nutrient-sensitive hYS genes differ from non-nutrient-sensitive hYS genes

Highly expressed nutrient-sensitive hYS genes were involved in 10 molecular functions, 15 biological processes, and 21 protein classes (Figure 3A-C). Nearly half (48%) of highly expressed nutrient-sensitive hYS genes were metabolite interconversion enzymes. The top molecular functions for nutrient-sensitive hYS genes were transferase, catalytic, and oxidoreductase activity, and the top biological processes were cellular and metabolic processes.

**Figure 3.**
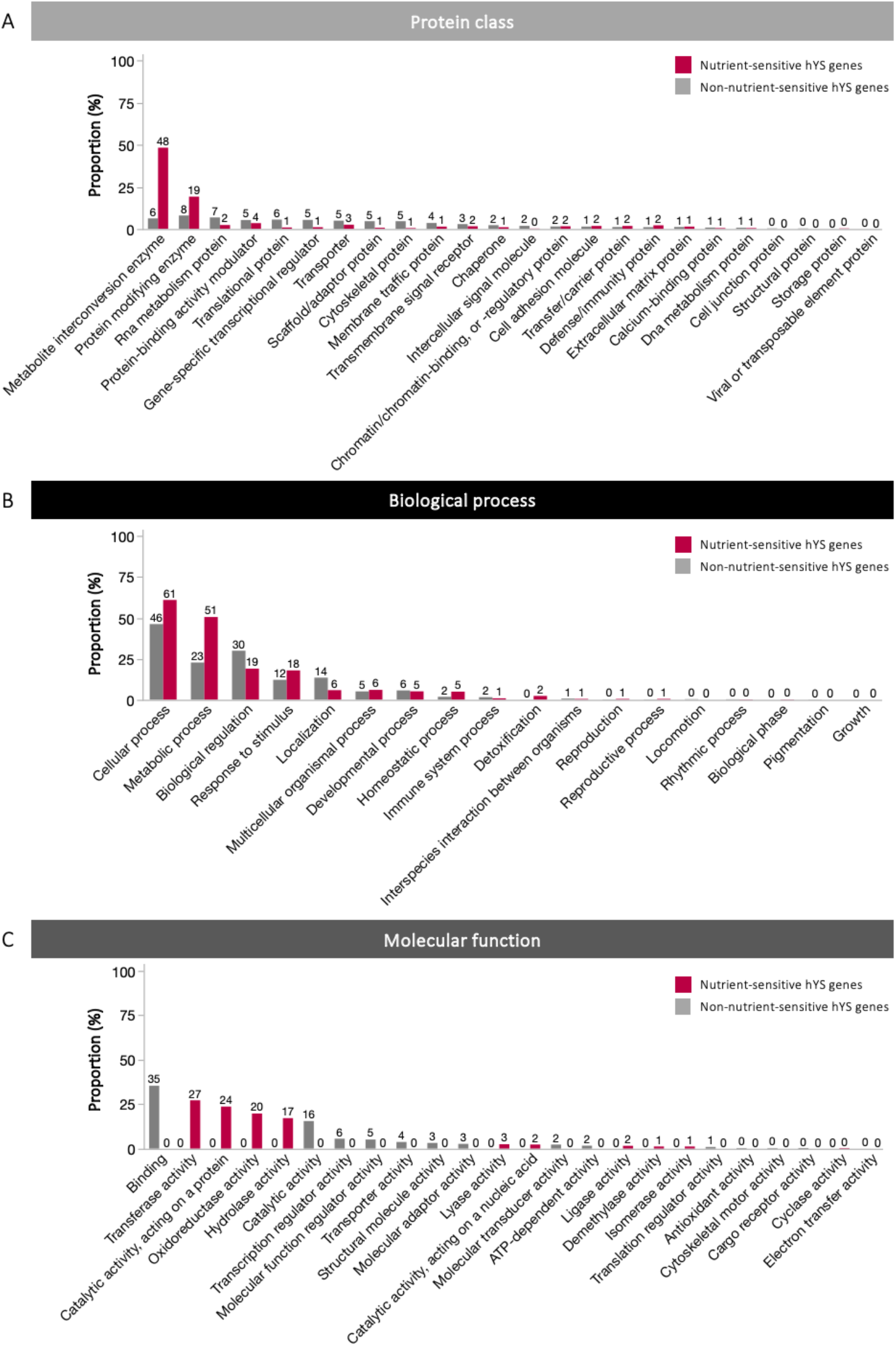
Functional analysis of highly expressed hYS genes. (A) Protein classes, (B), biological processes, and (C) molecular functions of highly expressed hYS genes that are or are not nutrient-sensitive. The numbers above the bars are the proportion of highly expressed, nutrient-sensitive or non-nutrient-sensitive hYS genes that fell within that category.

Of the 109 pathways that involved >1 highly expressed nutrient-sensitive hYS gene, 26 involved >5 nutrient-sensitive hYS genes (Figure 4). The pathways with the most nutrient-sensitive hYS genes were the gonadotropin-releasing hormone receptor pathway (n=25 nutrient-sensitive hYS genes), integrin signalling pathway (n=19), and the CCKR signaling map pathway (n=18).

**Figure 4.**
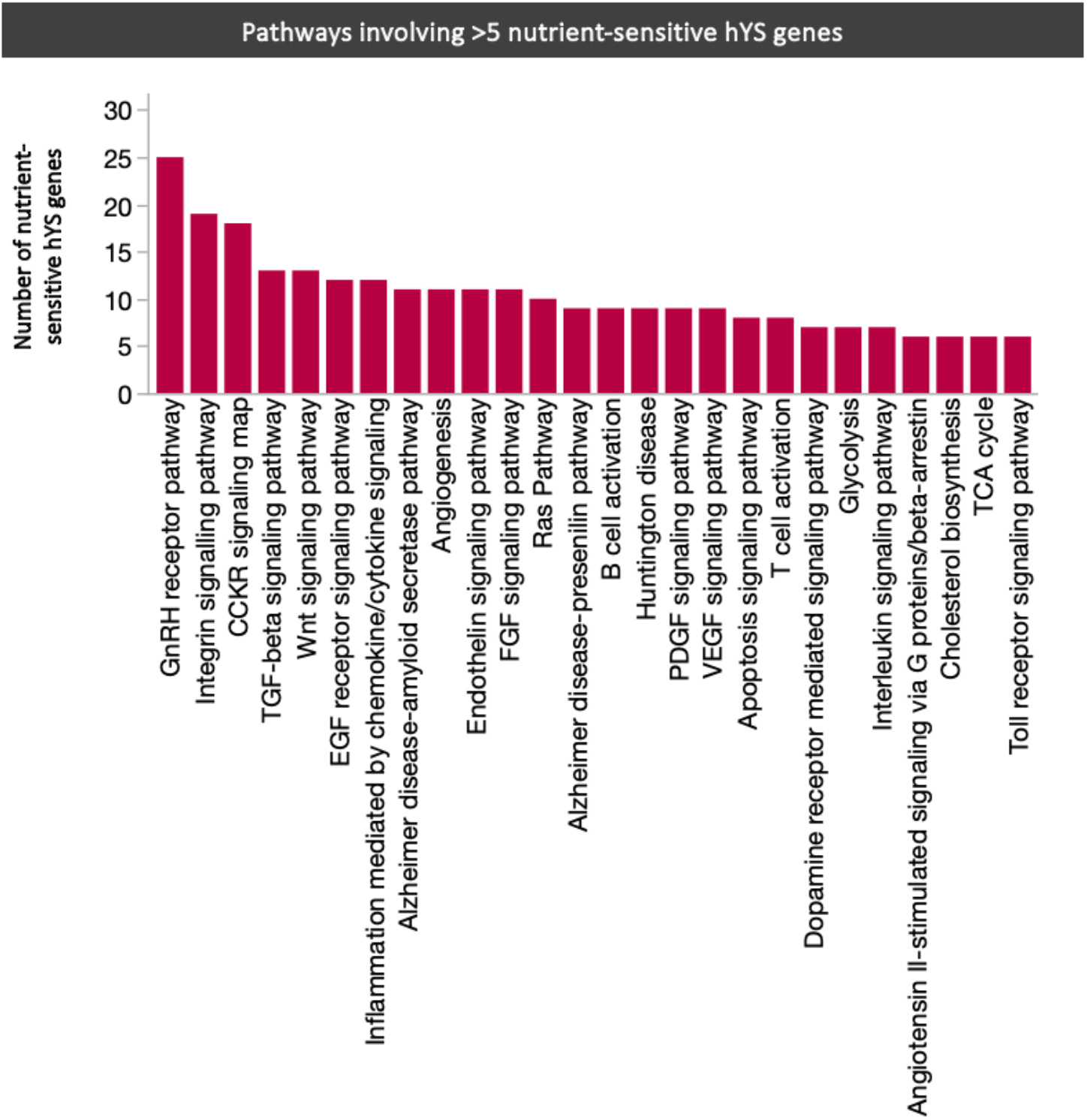
PANTHER pathways involving >5 nutrient-sensitive hYS genes. GnRH = gonadotropin-releasing hormone; TGF = Transforming growth factor; EGF = Epidermal growth factor; FGF = Fibroblast growth factor; VEGF = Vascular endothelial growth factor; PDGF = Platelet derived growth factor.

### Nutrient-sensitive TRs regulate highly expressed hYS genes

A total of 18 miRNAs were expressed in the hYS, of which, 16 were lowly expressed and two were highly expressed (MIR3916 and MIR24-2). MIR3916 was known to target one highly expressed, nutrient-sensitive hYS gene (Superoxide Dismutase 2 [SOD2]) which has a manganese cofactor. MIR24-2 was not known to target any highly expressed, nutrient-sensitive hYS genes.

A total of 60 TRs predicted to regulate highly expressed hYS genes were identified, of which four TRs were nutrient-sensitive and collectively regulated 1127 highly expressed hYS genes (Figure 5 and Supplementary Table S2). The nutrient-sensitive TRs included YY1 Transcription factor (YY1; sensitive to Zn; regulates 857 hYS genes), TATA-Box Binding Protein Associated Factor 1 (TAF1; sensitive to Mg; regulates 557 genes), Tumor Protein P53 (TP53; sensitive to Zn; regulates 218 genes), and RNA Polymerase II Subunit A (POLR2A; sensitive to Zn and Mg; regulates 857 genes).

**Figure 5.**
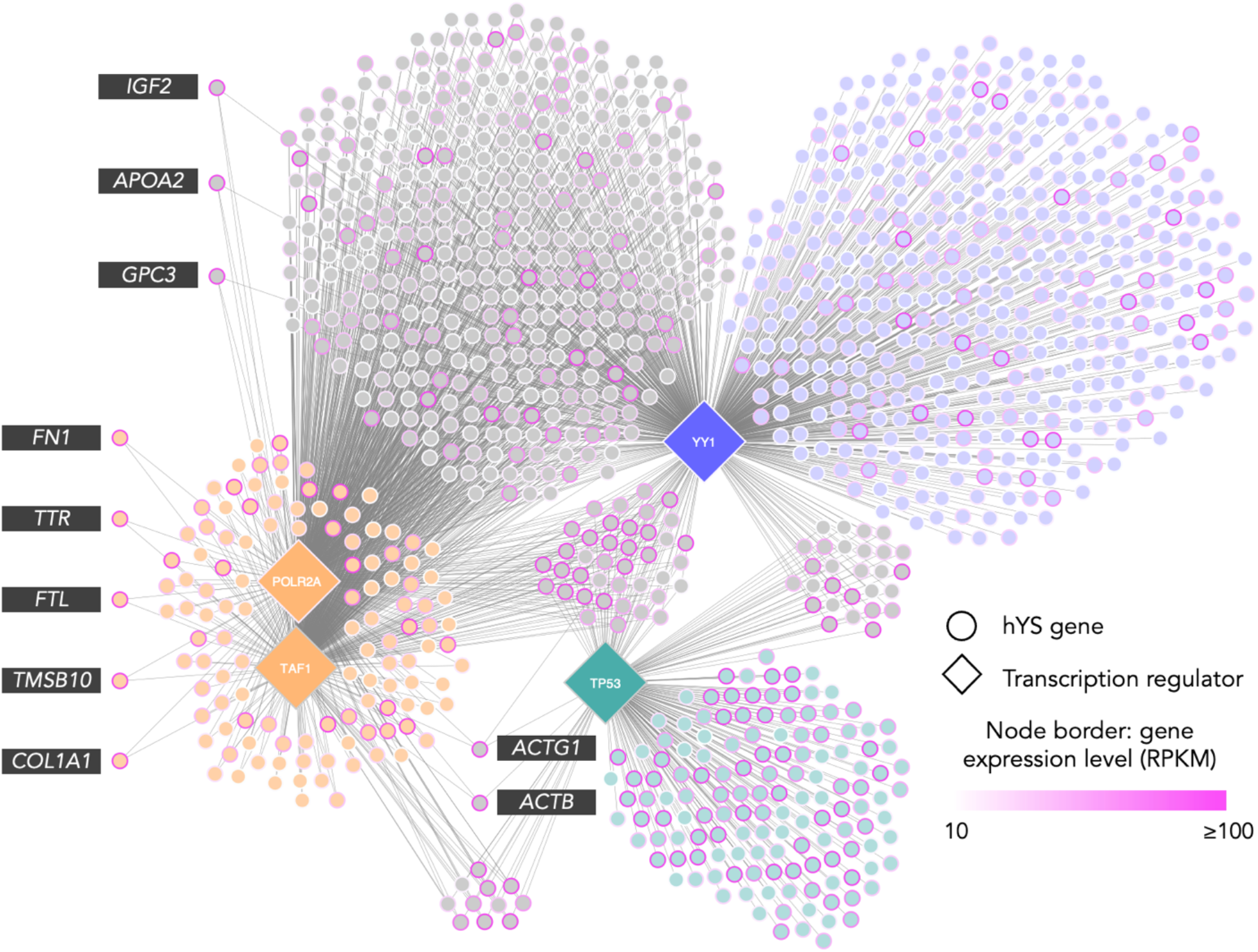
Nutrient-sensitive transcription regulators of hYS genes. Nutrient-sensitive transcription regulators (diamond nodes; n=4) connected to hYS gene targets (circle nodes; n=1127). Node border colour = gene expression in RPKM (reads per kilobase of transcript), per million mapped reads. The top 10 most highly expressed hYS gene targets are labelled. *ACTB* = Actin Beta; *ACTG1* = Actin Gamma 1; *APOA2* = Apolipoprotein A2; *COL1A1* = Collagen Type I Alpha 1; *FN1* = Fibronectin 1; *FTL* = Ferritin Light Chain; *GPC3* = Glypican 3; *IGF2* = Insulin Like Growth Factor 2; *TMSB10* = Thymosin Beta 10; *TTR* = Transthyretin.

Functional analysis revealed that hYS genes targeted by TP53 were largely translational proteins, metabolite interconversion enzymes, and RNA metabolism and cytoskeletal proteins, involved in binding, catalytic activity, and structural molecule activity (Supplementary Figure S4A). The majority of TAF1 and POLR2A gene targets in hYS were cytoskeletal proteins, gene-specific transcriptional regulators, and protein modifying- and metabolite interconversion enzymes involved in binding (Supplementary Figure S4B). The top protein class for YY1 hYS gene targets was RNA metabolism proteins, followed by protein modifying enzymes, and most of the genes were involved in binding (Supplementary Figure S4C). The most common biological processes for hYS gene targets of all four TRs were cellular and metabolic processes and biological regulation (Supplementary Figure S4A-C).

### Iron and zinc transporters are the most abundant micronutrient transporters in the hYS

There were 41 highly expressed hYS genes that encoded a micronutrient transporter protein. Most abundant were iron transporters (n=12) and zinc transporters (n=6), followed by chloride (n=5), vitamin A (n=3), calcium (n=3), phosphate (n=2), magnesium (n=2), manganese (n=1), copper (n=1), selenium (n=1), heme (n=1), and vitamins B9 (n=1), B7 (n=1), B1 (n=1), and B12 (n=1; Figure 6). One additional highly expressed nutrient transporter in the hYS, scavenger receptor class B member 1 (SCARB1), is a plasma membrane protein that binds high density lipoprotein cholesterol [77].

**Figure 6.**
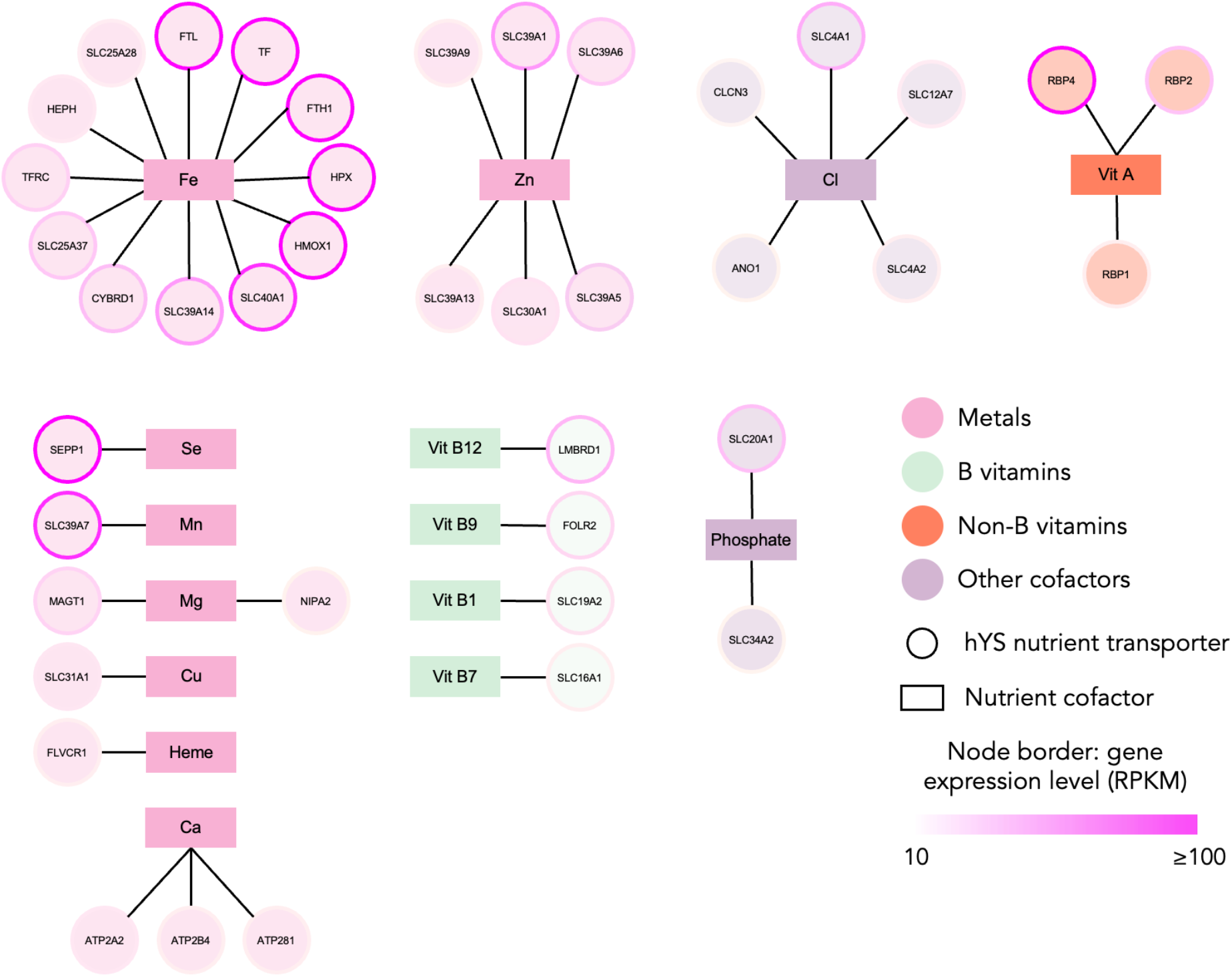
Highly expressed hYS genes encoding for nutrient transporters. hYS nutrient transporters (circle nodes) connected to the nutrient that they transport (rectangle nodes) via lines (edges). Node border colour = gene expression in RPKM (reads per kilobase of transcript), per million mapped reads. Fe = iron; Zn = zinc; Cl = chloride; Mg; magnesium; Mn = manganese; Cu = copper; Se = selenium; Ca = calcium.

Thirteen of the 41 micronutrient transporters identified were nutrient-sensitive proteins, including three calcium transporters with a magnesium cofactor and one with a calcium cofactor, six iron transporters with iron, heme, copper, and/or vitamin B3 cofactors, one folate transporter with a folate cofactor and two vitamin A transporters with a vitamin B6 cofactor, and one chloride transporter with a chloride cofactor (Supplementary Table S3).

### hYS genes with nutrient cofactors, namely calcium, encode proteins found in CF

We next determined whether, and which, nutrient-sensitive hYS genes are expressed as proteins in the human CF. Twenty-five of the 248 proteins known to be expressed in human CF were encoded by nutrient-sensitive genes that were also highly expressed in hYS. The hYS genes encoding proteins found in CF were sensitive to calcium (n=15 genes), zinc (n=5), copper (n=4), glutathione (n=2), vitamin B5 (n=1), iron (n=1), and orthoquinone (n=1; Table 1).

**Table 1.** Function of genes expressed in the human yolk sac that encode a protein found in human coelomic fluid.

| Gene | Symbol | Cofactor | Function |
| --- | --- | --- | --- |
| Albumin | *ALB* | Cu, Zn | Encodes for albumin, the most abundant protein in human blood and functions to plasma osmotic pressure. It is also responsible for binding 45% of free calcium and magnesium. |
| Diazepam binding inhibitor, acyl-CoA binding protein | *DBI* | Vitamin B5 | Encodes a protein regulated by hormones that functions in fatty acid metabolism, in specific it binds medium- and long-chain acyl-CoA esters with very high affinity. It also acts as a neuropeptide by modulating the action of the GABA receptor. |
| Collagen type I alpha 1 chain | *COL1A1* | Ca | Encodes a member of the group I collagens (fibrillar forming collagen). |
| Alpha fetoprotein | *AFP* | Metal | Encodes alpha-fetoprotein, a protein known to be expressed in the yolk sac during early development and binds to copper, nickel, and fatty acids, and bilirubin less well than, serum albumin. |
| Heparan sulfate proteoglycan 2 | *HSPG2* | Ca | Encodes an essential component of the basement membrane. It plays essential roles in vascularisation, heart development, avascular cartilage development, and has anti-angiogenic effects. Since it is a proteoglycan on the cell surface it is known to bind to the ECM and cell-surface molecules. |
| Collagen type III alpha 1 chain | *COL3A1* | Ca | Encodes a member of the group III collagens, which are found most in soft connective tissue along with type 1 collagens. |
| Collagen type I alpha 2 chain | *COL1A2* | Ca | Encodes a member of the group I collagens (fibrillar forming collagen). |
| Transferrin | *TF* | Fe | Encodes an iron binding protein which is responsible for transport of iron from sites of absorption, and heme degradation to storage and iron utilization sites. |
| Glutathione peroxidase 3 | *GPX3* | GSH | Encodes a protein which protects a cell from oxidative damage by catalyzing the reduction of hydrogen peroxide, lipid peroxides and organic hydroperoxide, by glutathione. |
| Gelsolin | *GSN* | Ca | Encodes an actin-modulation protein that binds to monomers and prevents them from creating polymer filaments. |
| Collagen type XVIII alpha 1 chain | *COL18A1* | Ca | Encodes the alpha chain of type XVIII collagen, which inhibits endothelial cell proliferation and angiogenesis (by binding heparan sulfate proteoglycans involved in growth factor signaling). Likely plays major role in determining the retinal structure as well as in the closure of the neural tube. |
| Ectonucleotide pyrophosphatase/phosphodiesterase 2 | *ENPP2* | Ca, Zn | Encodes the primary enzyme responsible for generating lysophosphatidic acid (LPA). Also has pro-angiogenic functions and promotes tumor motility. |
| Collagen type V alpha 2 chain | *COL5A2* | Ca | Encodes a type V collagen chain which is a member of group I collagens (fibrillar forming collagen). Which binds heparan sulfate, thrombospondin, heparin, and insulin. |
| Prostaglandin D2 synthase | *PTGDS* | GSH | Encodes an enzyme which creates a prostaglandin involved in smooth muscle contraction/relaxation and which is a potent inhibitor of platelet aggregation. |
| Collectin subfamily member 12 | *COLEC12* | Ca | Encodes a C-type lectin which mediates the recognition, internalization and degradation of oxidatively modified low-density lipoprotein (oxLDL) by vascular endothelial cells. |
| Coagulation factor 5 | *F5* | Ca, Cu | Encodes an essential cofactor for prothrombinase activity of factor Xa that results in the activation of prothrombin to thrombin. |
| Cadherin 1 | *CDH1* | Ca | Encodes a cadherin protein, which is involved in cell-cell adhesion and is calcium dependent. |
| TIMP metalloproteinase inhibitor 2 | *TIMP2* | Zn | Encodes a metalloproteinase (MMP; typically known to break down ECM components) inhibitor forming irreversibly inactivating them by binding to their catalytic zinc cofactor. Has anti-angiogenic properties. |
| ADAM metalloproteinase with thrombospondin type 1 motif 1 | *ADAMTS1* | Ca, Zn | Encodes an active metalloproteinase that may be associated with inflammatory responses, also has anti-angiogenic properties. |
| Cadherin 2 | *CDH2* | Ca | Encodes a cadherin protein, which is involved in cell-cell adhesion and is calcium dependent. |
| Mannosidase alpha class 1A member 1 | *MAN1A1* | Ca | Encodes a mannosidase involved in the maturation of Asn-linked oligosaccharides |
| Lysyl oxidase | *LOX* | Cu, Orthoquinone | Encodes a protein involved in post-translational oxidative deamination of peptidyl lysine residues in precursors to fibrous collagen and elastin. Plays a role in tumor suppression and aortic wall architecture. |
| Bone morphogenetic protein 1 | *BMP1* | Zn | Encodes a metalloproteinase that plays key roles in regulating the formation of the extracellular matrix (ECM). An isoform plays a role in bone repair by acting as a cofactor for BMP7. |
| Superoxide dismutase 1 | *SOD1* | Cu | Encodes an enzyme which destroys radicals produced in cells, and which would be toxic if not cleared. |
| Collagen type V alpha 1 chain | *COL5A1* | Ca | Encodes an alpha chain for the type V collagen, which is a fibrillar forming collagen. It is a common component of minor connective tissue. It also DNA, heparan sulfate, thrombospondin, heparin, and insulin |

Ca = calcium; Zn = zinc; Cu = copper; GSH = glutathione; Fe = iron; Mg = magnesium; K = potassium.
All key information on gene (protein) functions and processes were obtained from genecards.org [77].

Functional analysis revealed nine PANTHER pathways involving >1 of the nutrient-sensitive hYS gene-translated CF proteins. The pathways involving the most nutrient-sensitive hYS genes that were expressed as proteins in CF were integrin signalling pathway (n=5 hYS genes), cadherin signaling pathway (n=2), Wnt signaling pathway (n=2), and blood coagulation pathway (n=2). The other six pathways each involved one hYS gene: angiogenesis pathway, Alzheimer disease-presenilin pathway, TGF-beta signaling pathway, CCKR signaling map pathway, and the FAS signaling pathway.

### Nutrient-sensitive hYS genes associate with perinatal pathologies with embryonic origins

Last, we identified and described nutrient-sensitive hYS genes that are associated with perinatal pathologies with embryonic origins. A total of 162 hYS gene-phenotype associations were identified (121 established in humans, 41 established in mouse models), involving 117 unique nutrient-sensitive hYS genes and 10 perinatal pathologies (Figure 7A). The highest number of nutrient-sensitive hYS were associated with CHDs (n=54 hYS genes), followed by microcephaly (n=37), abnormal YS morphology (n=31), NTDs (n=13), spontaneous abortion (n=12), renal agenesis (n=6), exomphalos (n=4), gastroschisis (n=2), anorectal atresia (n=2), and orofacial clefts (n=1; Figure 7A and Supplementary Table S4).

**Figure 7.**
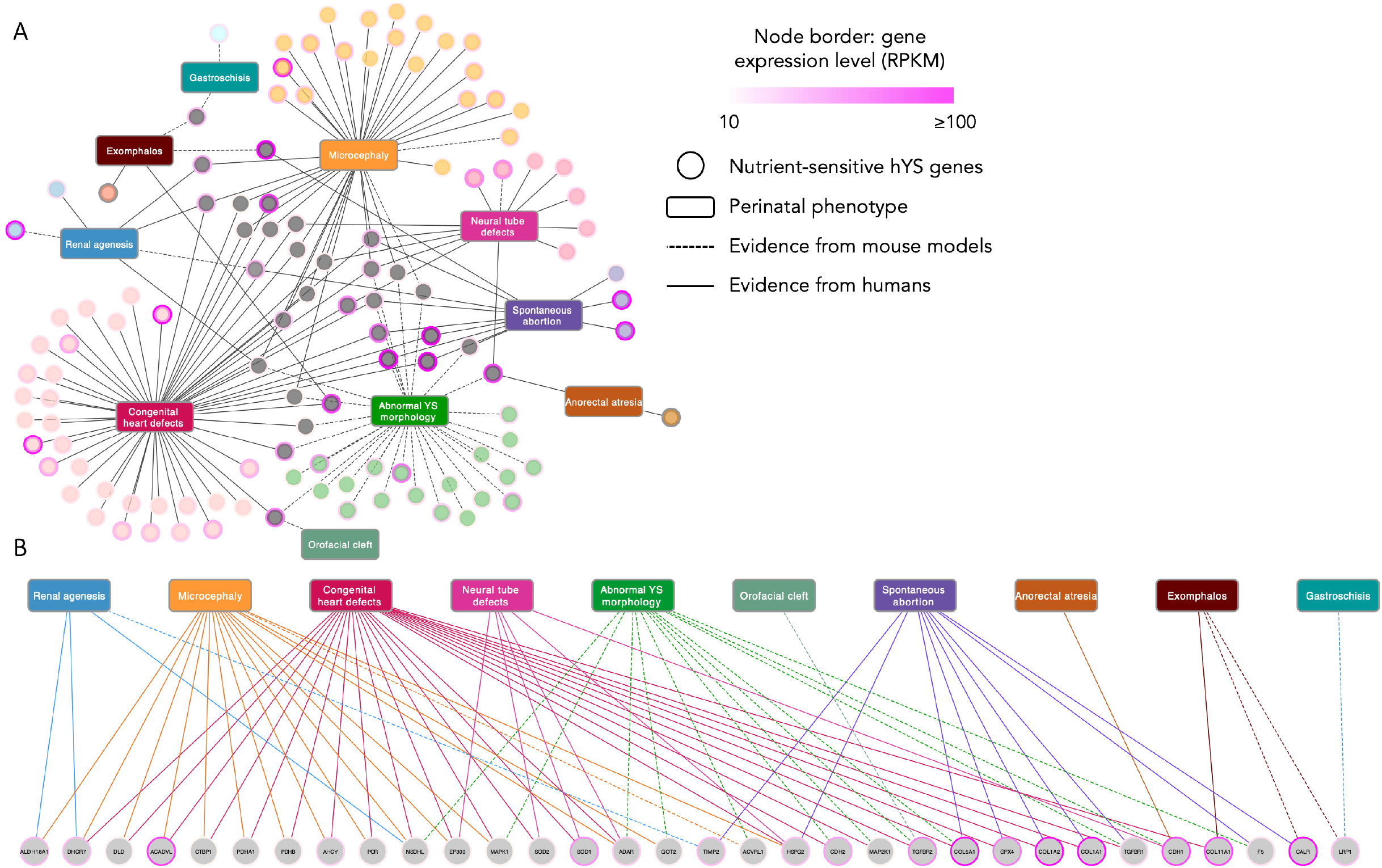
Adverse perinatal phenotypes with embryonic origins associate with highly expressed nutrient-sensitive hYS genes. (A) Connections between adverse perinatal phenotypes of embryonic origin and highly expressed nutrient-sensitive hYS genes (circle nodes; n=117). (B) Highly expressed hYS genes associated with >1 adverse perinatal phenotype. Genes are colour in coordination with associated phenotypes. Genes associated with >1 phenotype are grey (n=32). Dotted edges = mouse disease association. Full edges = human disease association. Node border colour = gene expression in RPKM (reads per kilobase of transcript), per million mapped reads.

Thirty-two of the 117 highly expressed nutrient-sensitive genes associated with >1 pathological perinatal phenotype (Figure 7B and Supplementary Table S5), for which the most abundant cofactors were calcium (n=10 hYS genes), B vitamins (n=9), zinc (n=3), and magnesium (n=5; Table 2). The functions of genes known to associate with >1 pathological perinatal phenotype included lipid metabolism (n=5 hYS genes; sensitive to vitamins B2 and B3, calcium, and CoQ), encoding for a collagen chain (n=4; sensitive to calcium), the clearance of free radicals (n=3; sensitive to copper, zinc, manganese, SAM, and GSH), cell cycle regulation (n=3; sensitive to magnesium, vitamin B5, and zinc), cell-cell adhesion (n=4; sensitive to calcium), vascular development (n=4; sensitive to magnesium, calcium, and metal cations), the tricarboxylic acid cycle (n=3; sensitive to potassium, magnesium, CoQ, and vitamin B5) and amino acid metabolism (n=2; sensitive to vitamin B2; Table 2 [78-95]).

**Table 2.** Function of highly expressed nutrient-sensitive genes expressed in the human yolk sac that are known to associate with >1 perinatal pathological phenotype.

| Gene Name | Gene Symbol | Nutrient Cofactor | Known Target/Function | Associated Perinatal Pathological Phenotypes |
| --- | --- | --- | --- | --- |
| Collagen type I alpha 1 chain | COL1A1 | Ca | Encodes an alpha 1 chain of collagen type I, which is a fibril-forming collagen and is found in most connective tissues (highly expressed in bone, tendon, cornea, and dermis). Deletion of COL1A1 in mice offspring results in arrested embryonic development and embryonic lethality around embryonic day 6–7 (E6-7). | Congenital heart defects<br>Spontaneous abortion |
| Collagen type XI alpha 1 chain | COL11A1 | Ca | Encodes an alpha 1 chain of collagen type XI, which is a minor fibrillar forming collagen and may be involved in regulating the lateral growth of collagen II fibrils. Required for embryonic fibril formation and endochondral ossification in the embryo. | Congenital heart defects<br>Exomphalos |
| Coagulation factor V | F5 | Ca, Cu | Encodes coagulation factor V, which binds to an activation factor released by thrombin during coagulation. Once activated, this protein is a heavy and light chain held together by calcium ions and along with activated coagulation factor X, activates the transition of prothrombin to thrombin, aiding in the clotting cascade process. Activated protein C (APC) inactivates coagulation factor V, slowing the clotting process. Needed for yolk sac vasculature development, when deleted it leads to embryonic death at E9-10 in mice, if insufficient the increase of thrombosis risk. | Abnormal yolk sac morphology<br>Spontaneous abortion |
| Collagen type I alpha 2 chain | COL1A2 | Ca | Encodes an alpha 2 chain of collagen type I, which is a fibril-forming collagen and is found in most connective tissues (highly expressed in bone, tendon, cornea, and dermis). | Congenital heart defects<br>Spontaneous abortion |
| Glutathione peroxidase 4 | GPX4 | GSH, SAM | Encodes a peroxidase enzyme involved in reducing hydrogen peroxide, organic hydroperoxides and lipid hydroperoxides. Meaning it plays an important role in protecting the cell from oxidative damage. Regulates dorsal organizer formation in zebrafish embryos via inhibition of Wnt/β-catenin signals. | Congenital heart defects<br>Spontaneous abortion |
| Collagen type V alpha 1 chain | COL5A1 | Ca | Encodes an alpha 1 chain of collagen type V, which is a fibril-forming collagen expressed ubiquitously in all soft tissue. Needed for fibril assembly in embryonic mesenchyme, lethal if deleted in mice. | Congenital heart defects<br>Abnormal yolk sac morphology<br>Spontaneous abortion |
| Transforming growth factor beta receptor 1 | TGFBR1 | Mg | Encodes transforming growth factor (TGF) beta receptor 1, which forms a heteromeric complex with TGF beta receptor 2 when bound to TGF-beta. Thus, TGFBR1 is important in initiating the TGF-beta mediates physiological responses, including cell cycle arrest in epithelial and hematopoietic cells, control of mesenchymal cell proliferation and differentiation, wound healing, extracellular matrix production, immunosuppression, and carcinogenesis. Mutants cause issues in SMC defects and lowered contractile protein transcripts. | Congenital heart defects<br>Abnormal yolk sac morphology<br>Spontaneous abortion |
| Cadherin 2 | CDH2 | Ca | Encodes a classical cadherin molecule, which is a calcium-dependent adhesion protein. It plays a role in cell-cell junction adhesion between neural crest stem cells (NCS) and may be involved in a neuronal recognition system and/or regulating dendritic spine density in the hippocampus. Regulator of stem cell fate/specification. | Congenital heart defects<br>Abnormal yolk sac morphology |
| Calreticulin | CALR | Ca | Encodes a chaperone protein primarily found mainly in the endoplasmic reticulum (ER) and is involved in cell adhesion, protein folding, and calcium homeostasis. In mice, mutations result in thrombocythemia (ET) and primary myelofibrosis (PMF), implicating dysregulation in blood cell differentiation and proliferation. | Exomphalos<br>Spontaneous abortion |
| Ldl receptor related protein 1 | LRP1 | Ca | Encodes a low-density lipid (LDL) receptor, is thus involved in lipid homeostasis and acts as an endocytic and phagocytic receptor for apoptotic cells. It is also required for embryogenesis and plays roles in the clearance of molecules such as amyloid precursors, activated LRPAP1 (alpha 2-macroglobulin), and plasminogen activators and inhibitors. | Exomphalos<br>Gastroschisis |
| Transforming growth factor beta receptor 2 | *TGFBR2* | Mg | Encodes transforming growth factor (TGF) beta receptor 2, which forms a heteromeric complex with TGF beta receptor 1 when bound to TGF-beta. Thus, TGFBR1 is important in initiating the TGF-beta mediates physiological responses, including cell cycle arrest in epithelial and hematopoietic cells, control of mesenchymal cell proliferation and differentiation, wound healing, extracellular matrix production, immunosuppression, and carcinogenesis. | Congenital heart defects<br>Abnormal yolk sac morphology<br>Orofacial cleft |
| Cadherin 1 | *CDH1* | Ca | Encodes a classical cadherin molecule, which is a calcium-dependent adhesion protein. It also plays a role in regulating epithelial cell's ability to be motile, proliferate, and bind other cells. | Neural tube defects<br>Abnormal yolk sac morphology<br>Anorectal atresia |
| Mitogen-activated protein kinase kinase 1 | *MAP2K1* | Mg | Encodes a mitogen-activated protein (MAP) kinase. MAP kinases have widely varying roles in propagating biochemical signals, and MAP2K1 can lead to varying different biochemical pathways depending on the upstream signal. Required for normal syncytiotrophoblast development during placentation. | Congenital heart defects<br>Abnormal yolk sac morphology |
| Heparan sulfate proteoglycan 2 | *HSPG2* | Ca | Encodes a perlecan protein, which plays a role in maintaining the endothelial cell junctions via it strengthening the vasculature extracellular matrix (ECM). Component of perlecan which is needed for ECM structures, and required for cephalic and cartilage development, mice with deleted gene die at E10.5. | Spontaneous abortion<br>Congenital heart defects<br>Microcephaly<br>Neural tube defects |
| Superoxide dismutase 1 | *SOD1* | Cu, Zn | Encodes a superoxidase dismutase enzyme that destroys free radicals once bound to copper and zinc. Required for Wnt-dependent cell proliferation. | Congenital heart defects<br>Neural tube defects |
| Superoxide dismutase 2 | *SOD2* | Mn | Encodes a superoxidase dismutase enzyme that destroys free ion radicals once bound to one magnesium ion per subunit. Required for Wnt-dependent cell proliferation. | Congenital heart defects<br>Neural tube defects |
| Adenosine deaminase RNA specific | *ADAR* | Vitamin C | Encodes an enzyme involved in RNA editing, specifically in the deamination of adenosine to inosine. | Congenital heart defects<br>Microcephaly<br>Abnormal yolk sac morphology<br>Neural tube defects |
| Activin a receptor like type 1 | *ACVRL1* | Metal cation | Encodes a cell surface receptor for the TGF-beta family of proteins. This is a type one receptor and thus is important for normal blood vessel development. Encodes a ALK1 ligand which is essential in promoting vascular development and maintenance. | Microcephaly<br>Abnormal yolk sac morphology |
| Glutamic-oxaloacetic transaminase 2 | *GOT2* | Vitamin B5 | Encodes glutamic-oxaloacetic transaminase, which is an enzyme important in amino acid metabolism, urea cycle, and tricarboxylic acid cycle. | Microcephaly<br>Abnormal yolk sac morphology |
| Mitogen-activated protein kinase 1 | *MAPK1* | Mg | Encodes a mitogen-activated protein (MAP) kinase 1. MAP kinases have widely varying roles in propagating biochemical signals, and MAPK1 can lead to varying different biochemical pathways depending on the upstream signal. Acts as a mediator for prostaglandins to promote angiogenesis and embryo-maternal interactions during implantation. | Congenital heart defects<br>Microcephaly<br>Abnormal yolk sac morphology |
| E1a binding protein p300 | *EP300* | Vitamin B5, Zn | Encodes a protein involved in chromatin remodelling via its histone acetylation role. It has also been shown to have a role in cell proliferation and differentiation. | Congenital heart defects<br>Microcephaly<br>Neural tube defects |
| NAD(P) dependent steroid dehydrogenase-like | *NSDHL* | Vitamin B3 | Encodes a protein localized in the endoplasmic reticulum (ER) and is involved in cholesterol metabolism. NSDHL deficiency in mice lead to maldevelopment of placenta through altered Hedgehog pathway. | Renal agenesis<br>Congenital heart defects<br>Microcephaly<br>Abnormal yolk sac morphology |
| Dihydrolipoamide dehydrogenase | *DLD* | Vitamin B2, Vitamin B3 | Encodes a member of the class-I pyridine nucleotide-disulfide oxidoreductase family, which can function as a dehydrogenase and a protease. It has been shown to be involved in three alpha-ketoacid dehydrogenase complexes. | Congenital heart defects<br>Microcephaly |
| Pyruvate dehydrogenase E1 subunit alpha 1 | *PDHA1* | K, Mg, CoQ | Encodes an alpha 1 subunit to pyruvate dehydrogenase (PDH), which is a key enzyme in the tricarboxylic acid cycle, and thus cell energy metabolism. | Congenital heart defects<br>Microcephaly |
| Cytochrome P450 oxidoreductase | *POR* | Vitamin B2, Vitamin B3 | Encodes an oxidoreductase enzyme within the ER membrane and is important in catalyzing cytochrome P450-dependent reactions, like the metabolism of steroid hormones, drugs, and xenobiotics. Required for vascularization in early mice embryo, and in molecular patterning of the brain, abdominal/caudal region and limbs. | Congenital heart defects<br>Microcephaly |
| C-terminal binding protein 1 | *CTBP1* | Vitamin B3 | Encodes a corepressor protein that targets various transcription regulators. It also has dehydrogenase activity and is involved in controlling the ratio of stacked and tubular structures in the Golgi apparatus. Enhances endometrial epithelial cell's ability to attach. | Congenital heart defects<br>Microcephaly |
| Pyruvate dehydrogenase E1 subunit beta | *PDHB* | K, CoQ | Encodes a beta subunit to pyruvate dehydrogenase (PDH), which is a key enzyme in the tricarboxylic acid cycle, and thus cell energy metabolism. | Congenital heart defects<br>Microcephaly |
| Acyl-CoA Dehydrogenase Very Long Chain | *ACADVL* | CoQ | Encodes very long chain to the acetyl-CoA dehydrogenase enzyme, which binds to the inner mitochondrial membrane and catalyzes the first step in the mitochondrial fatty acid beta-oxidation. Downregulated in non- decidualized endometrial stromal cell samples. | Congenital heart defects<br>Microcephaly |
| Adenosylhomocysteinase | *AHCY* | Vitamin B2 | Encodes an adenosylhomocysteinase enzyme which has a hydrolysis role leading to the production of adenosine (local transmethylation reaction). It also acts as a competitive inhibitor of S-adenosyl-L-methionine-dependent methyl transferase reactions. | Congenital heart defects<br>Microcephaly |
| 7-dehydrocholesterol reductase | *DHCR7* | Vitamin B3 | Encodes a reductase enzyme which reduces the 7<sup>th</sup> double bond in B ring of sterols and catalyzes the formation of cholesterol from 7-dehydrocholesterol. Important for cholesterol synthesis needed during early brain development. | Renal agenesis<br>Congenital heart defects<br>Microcephaly |
| Aldehyde dehydrogenase 18 family member A1 | *ALDH18A1* | Vitamin B2 | Encodes an aldehyde dehydrogenase enzyme which performs intermediate steps prior to the synthesis of proline, ornithine, and arginine. | Renal agenesis<br>Microcephaly |
| TIMP metalloproteinase inhibitor 2 | *TIMP2* | Zn | A natural inhibitor of matrix metalloproteinases, enzymes involved in extracellular matrix degradation, this protein uniquely suppresses endothelial cell proliferation; may be key in maintaining tissue homeostasis by inhibiting protease activity and controlling tissue response to angiogenic factors. | Spontaneous abortion<br>Renal agenesis |

Information on gene (protein) functions and processes were obtained from genecards.org or published literature [77].
logFC = log fold change; CoQ = Coenzyme Q10; NAD = Nicotinamide Adenine Dinucleotide; GSH = Glutathione; SAM = S-Adenosyl methionine; Ca = calcium, Cu = copper, Mg = magnesium, Zn = zinc, K = potassium.

## Discussion

Here we used a nutrient-focused analysis approach to gain novel insights Here we used a nutrient-focused analysis approach to gain novel insights on nutrient-sensitive biosynthesis, signalling, and transport roles of the hYS in uncomplicated pregnancies. We found that genes with B vitamin and metal cofactors are highly expressed in the hYS, and that iron and zinc transporters were the most abundant nutrient transporters in the hYS. Our finding that hYS genes with nutrient cofactors, namely calcium, encode proteins that are also found in CF supports that nutrient-sensitive gene networks in the hYS may interact (in)directly with micronutrients in the CF to regulate embryonic development. Finally, we characterised hYS genes with metal and vitamin B6 cofactors that associate with multiple adverse perinatal phenotypes with embryonic origins. Collectively, these results contribute new understandings of micronutrient-gene interactions in hYS tissue, hYS physiology, and the pathogenesis of nutrient-sensitive pathologies that originate during embryogenesis.

It is known that a primary function of the hYS is to mediate embryonic nutrition and protection [1, 5, 6], and here we have described how the hYS interacts with nutrients in at least two ways: firstly, highly expressed genes in the hYS are sensitive to the bioavailability of several nutrients, and secondly, the hYS expresses multiple nutrient transporters, ensuring sufficient nutrient exchange between the CF and the developing embryo. hYS genes with nutrient cofactors, namely calcium, were also found in the CF as proteins, suggesting that nutrient-sensitive hYS functions may be influenced by micronutrient bioavailability and gene and protein signaling in the adjacent CF. Nutrient-sensitive genes/proteins expressed in both the hYS and CF are involved in the regulation of several key processes essential for embryonic development, including integrin, cadherin, Wnt, and TGF-β signaling, which support embryonic epithelial morphogenesis [96],[97], macrophage maturation [98], gastrulation [99], cell differentiation, migration and organogenesis [100]. Notably, nutrient-sensitive hYS genes were more likely to be involved in biosynthesis and metabolism-related processes than non-nutrient-sensitive hYS genes, reflecting the well-established role of nutrients in intermediary metabolism [101]. In line with this, we also found that six of the top 10 most highly expressed nutrient-sensitive hYS genes were encoded by the mitochondria, which plays a central role in intermediary metabolism by acting as the main centre of energy production and metabolic regulation [102]. Collectively, our finding that several highly expressed, nutrient-sensitive hYS genes are involved in these key pathways and developmental processes highlights the importance of nutrient-gene interactions in supporting hYS development and functions in uncomplicated pregnancies.

We also found that zinc and iron transporters were the most abundant nutrient transporters in the hYS, which may suggest that transport of these nutrients to the developing embryo is prioritized, consistent with the known importance of these nutrients for embryogenesis. Iron is essential for hematopoiesis [103] and zinc, a regulator of gene expression, is required to support the high rates of DNA synthesis and cell proliferation that occur during embryonic development [104, 105]. Zinc and B vitamins (namely vitamin B3) were also amongst the most common nutrient cofactors of highly expressed hYS genes, and three of four nutrient-sensitive TRs in the hYS had zinc cofactors, illustrating the importance of zinc and B vitamins for supporting hYS function in early pregnancy. It is plausible that insufficient or excessive availability of nutrient cofactors required for transcriptional regulation could lead to an altered pattern of gene expression in hYS pathways critical for development [106, 107]. Notably, we have previously shown that in human amniocytes and human placental tissues collected later in pregnancy from fetuses with congenital anomalies, zinc- and B vitamin-sensitive genes and gene regulatory networks are prevalent among dysregulated genes [38, 44]. As there is some evidence to suggest that zinc may play a role in vitamin B3 metabolism via interactions with vitamin B6 [108, 109], these findings collectively highlight the need for further study of zinc- and B vitamin-gene interactions in embryonic and gestational tissues, to better understand key nutrient-sensitive mechanisms of both typical and pathological development.

While suboptimal nutrient bioavailability during embryonic and fetal development has been linked to the formation of congenital anomalies [25-34], the mechanisms underlying the relationship between nutrient insufficiency or excess and the increased risk of these anomalies remain poorly understood. By using a nutrient-focused approach to analyze known gene-phenotype associations in perinatal pathologies with embryonic origins, we identified several nutrient-sensitive hYS genes linked to 10 of the most common congenital anomalies. These findings support that the hYS plays a role in mediating nutrient-sensitive disease risk for the developing embryo, and that nutrient-sensitive hYS dysfunction may contribute to the development of common congenital anomalies. Of the perinatal pathologies assessed, CHDs had the highest number of nutrient-sensitive hYS gene-phenotype associations, followed by microcephaly and abnormal YS morphology. The higher number of associations identified could be partly because gene-phenotype associations for these conditions are more well-researched than others, rather than due to greater nutrient sensitivity. As the incidence of some of these congenital anomalies is increasing [110, 111], there is a need to further elucidate environment (including nutrient)-gene interactions that play a role in the disease etiology, to identify novel targets for prevention.

This study has several strengths and limitations that warrant discussion. First, this study describes the nutrient-sensitive genes and pathways in the hYS and CF transcriptomic and proteomic environments, through a re-analysis of datasets from separate cohorts. This prevents us from statistically comparing gene and protein expression levels in the hYS and CF, and from performing additional experiments to confirm gene and protein expression levels in these tissues. Further, although we described associations between nutrient-sensitive hYS genes and perinatal pathologies originating during embryogenesis, we did not have access to hYS samples from pregnancies with congenital anomalies. Therefore, it remains unclear whether or how the expression of these hYS genes may be altered in pregnancies affected by the perinatal pathologies discussed, and this should be a focus of future studies. However, the logistical and ethical challenges of obtaining hYS samples from uncomplicated pregnancies make these existing datasets particularly valuable for re-analysis. Other strengths of this study include the use of a nutrient-focused analysis approach to hYS transcriptomics and CF proteomics, and the application of a nutrient-focused lens to describe hYS-gene phenotype associations for 10 of the most common congenital anomalies with embryonic origins. These approaches provide a global view of micronutrient-gene interactions in hYS tissue from uncomplicated pregnancies and contribute to our understanding of the ways in which nutrient-sensitive hYS functions influence both typical and pathological development.

This study characterised the nutrient-sensitive hYS roles in biosynthesis, signaling, and transport that support embryonic development, both in uncomplicated pregnancies and in the context of nutrient-sensitive congenital anomalies that may arise, in part, due to hYS dysfunction. Our investigation into how nutrients interact with gene and protein signaling networks in the hYS and CF microenvironment increases our understanding of the impact of early-life nutritional exposures on embryonic development and may ultimately inform the identification of novel nutrient-sensitive targets for preventing adverse perinatal phenotypes originating during embryogenesis.

## Data Availability

This study was a secondary analysis of previously published datasets [8, 11, 53], which have been made available by the study authors in electronic supplementary materials at https://www.nature.com/articles/s41598-018-29384-9 (CF tissue proteomics) and https://www.nature.com/articles/srep19633 (nutrient/nutrient cofactor transporters), and in a public data repository at www.ebi.ac.uk/ena/data/view/PRJEB18767 (hYS and CF tissue transcriptomics and proteomics).

## Author contributions

Conceptualization, KLC, MW, JAP, and EB; Methodology, MW, JAP, KLC, EB; Formal analysis, JAP, MW; Investigation, JAP, MW, KLC; Data curation, JAP, MW; Writing—original draft preparation, MW, JAP, and KLC; Writing—review and editing, MW, JAP, KLC, and EB; Visualisation, JAP, MW, KLC; Supervision, KLC and MW; Funding, KLC. All authors have read and agreed to the published version of the manuscript.

## References

[1] L. M. Martinelli, A. Carucci, V. J. H. Payano, K. L. Connor, and E. Bloise, “Translational Comparison of the Human and Mouse Yolk Sac Development and Function,” Reproductive Sciences, 2022/02/08 2022, doi: 10.1007/s43032-022-00872-8.

[2] I. Goh et al., “Yolk sac cell atlas reveals multiorgan functions during human early development,” (in eng), Science, vol. 381, no. 6659, p. eadd7564, Aug 18 2023, doi: 10.1126/science.add7564.

[3] I. V. Kuzmina, “The yolk sac as the main organ in the early stages of animal embryonic development,” (in eng), Front Physiol, vol. 14, p. 1185286, 2023, doi: 10.3389/fphys.2023.1185286.

[4] L. S. Mamsen, C. B. Brøchner, A. G. Byskov, and K. Møllgard, “The migration and loss of human primordial germ stem cells from the hind gut epithelium towards the gonadal ridge,” (in eng), Int J Dev Biol, vol. 56, no. 10-12, pp. 771–8, 2012, doi: 10.1387/ijdb.120202lm.

[5] L. M. Martinelli et al., “Malaria in pregnancy regulates P-glycoprotein (P-gp/Abcb1a) and ABCA1 efflux transporters in the Mouse Visceral Yolk Sac,” (in eng), J Cell Mol Med, vol. 24, no. 18, pp. 10636–10647, Sep 2020, doi: 10.1111/jcmm.15682.

[6] L. M. Martinelli et al., “Breast cancer resistance protein (Bcrp/Abcg2) is selectively modulated by lipopolysaccharide (LPS) in the mouse yolk sac,” (in eng), Reprod Toxicol, vol. 98, pp. 82–91, Dec 2020, doi: 10.1016/j.reprotox.2020.09.001.

[7] C. J. Jones and E. Jauniaux, “Ultrastructure of the materno-embryonic interface in the first trimester of pregnancy,” (in eng), Micron, vol. 26, no. 2, pp. 145–73, 1995, doi: 10.1016/0968-4328(95)00002-l.

[8] D. Aiello et al., “Human coelomic fluid investigation: A MS-based analytical approach to prenatal screening,” Scientific Reports, vol. 8, no. 1, p. 10973, 2018/07/20 2018, doi: 10.1038/s41598-018-29384-9.

[9] C. Virgiliou et al., “Metabolic profile of human coelomic fluid,” (in eng), Bioanalysis, vol. 9, no. 1, pp. 37–51, Jan 2017, doi: 10.4155/bio-2016-0223.

[10] E. Jauniaux, B. Gulbis, D. Jurkovic, S. Campbell, W. P. Collins, and H. A. Ooms, “Relationship between protein concentrations in embryological fluids and maternal serum and yolk sac size during human early pregnancy,” (in eng), Hum Reprod, vol. 9, no. 1, pp. 161–6, Jan 1994, doi: 10.1093/oxfordjournals.humrep.a138308.

[11] T. Cindrova-Davies, E. Jauniaux, G. Elliot Michael, S. Gong, J. Burton Graham, and D. S. Charnock-Jones, “RNA-seq reveals conservation of function among the yolk sacs of human, mouse, and chicken,” Proceedings of the National Academy of Sciences, vol. 114, no. 24, pp. E4753–E4761, 2017/06/13 2017, doi: 10.1073/pnas.1702560114.

[12] A. Saeed, M. Hoekstra, M. O. Hoeke, J. Heegsma, and K. N. Faber, “The interrelationship between bile acid and vitamin A homeostasis,” Biochimica et Biophysica Acta (BBA) - Molecular and Cell Biology of Lipids, vol. 1862, no. 5, pp. 496–512, 2017/05/01/ 2017, doi: 10.1016/j.bbalip.2017.01.007.

[13] M. Aboelenain et al., “Pyridoxine supplementation during oocyte maturation improves the development and quality of bovine preimplantation embryos,” Theriogenology, vol. 91, pp. 127–133, 2017/03/15/ 2017, doi: 10.1016/j.theriogenology.2016.12.022.

[14] D. B. Dalto and J.-J. Matte, “Pyridoxine (Vitamin B6) and the Glutathione Peroxidase System; a Link between One-Carbon Metabolism and Antioxidation,” (in eng), Nutrients, vol. 9, no. 3, p. 189, 2017, doi: 10.3390/nu9030189.

[15] R. H. Finnell, G. M. Shaw, E. J. Lammer, and T. H. Rosenquist, “Gene–Nutrient Interactions: Importance of Folic Acid and Vitamin B12 During Early Embryogenesis,” Food and Nutrition Bulletin, vol. 29, no. 2_suppl1, pp. S86–S98, 2008/06/01 2008, doi: 10.1177/15648265080292S112.

[16] I. M. Kim, K. C. Norris, and J. N. Artaza, “Chapter Twelve - Vitamin D and Cardiac Differentiation,” in Vitamins & Hormones, vol. 100, G. Litwack Ed.: Academic Press, 2016, pp. 299–320.

[17] D. W. Nebert, W. L. Wikvall K Fau-Miller, and W. L. Miller, “Human cytochromes P450 in health and disease,” (in eng), no. 1471–2970 (Electronic).

[18] S. E. Juul, R. J. Derman, and M. Auerbach, “Perinatal Iron Deficiency: Implications for Mothers and Infants,” Neonatology, vol. 115, no. 3, pp. 269–274, 2019, doi: 10.1159/000495978.

[19] L. Gambling, Henriette S. Andersen, and Harry J. McArdle, “Iron and copper, and their interactions during development,” Biochemical Society Transactions, vol. 36, no. 6, pp. 1258–1261, 2008, doi: 10.1042/BST0361258.

[20] K. Grzeszczak, S. Kwiatkowski, and D. Kosik-Bogacka, “The Role of Fe, Zn, and Cu in Pregnancy,” (in eng), Biomolecules, vol. 10, no. 8, p. 1176, 2020, doi: 10.3390/biom10081176.

[21] Y. Komiya, L.-T. Su, H.-C. Chen, R. Habas, and L. W. Runnels, “Magnesium and embryonic development,” (in eng), Magnesium research, vol. 27, no. 1, pp. 1–8, Jan-Mar 2014, doi: 10.1684/mrh.2014.0356.

[22] Y. Komiya and L. W. Runnels, “TRPM channels and magnesium in early embryonic development,” (in eng), The International journal of developmental biology, vol. 59, no. 7-9, pp. 281–288, 2015, doi: 10.1387/ijdb.150196lr.

[23] S. Moradan and M. Forouzeshfar, “Are abnormal yolk sac characteristics important factors in abortion rates?,” (in eng), Int J Fertil Steril, vol. 6, no. 2, pp. 127–30, Jul 2012.

[24] S. Tan et al., “Abnormal sonographic appearances of the yolk sac: which can be associated with adverse perinatal outcome?,” (in eng), Med Ultrason, vol. 16, no. 1, pp. 15–20, Mar 2014, doi: 10.11152/mu.2014.2066.161.st1gt2.

[25] R. H. Finnell et al., “Gene Environment Interactions in the Etiology of Neural Tube Defects,” (in eng), Front Genet, vol. 12, p. 659612, 2021, doi: 10.3389/fgene.2021.659612.

[26] N. Hovdenak and K. Haram, “Influence of mineral and vitamin supplements on pregnancy outcome,” (in eng), Eur J Obstet Gynecol Reprod Biol, vol. 164, no. 2, pp. 127–32, Oct 2012, doi: 10.1016/j.ejogrb.2012.06.020.

[27] K. Nasri et al., “Maternal 25-hydroxyvitamin D level and the occurrence of neural tube defects in Tunisia,” International Journal of Gynecology & Obstetrics, vol. 134, no. 2, pp. 131–134, 2016/08/01/ 2016, doi: 10.1016/j.ijgo.2016.01.014.

[28] D. Dilli et al., “Maternal and neonatal micronutrient levels in newborns with CHD,” Cardiology in the Young, vol. 28, no. 4, pp. 523–529, 2018, doi: 10.1017/S1047951117002372.

[29] P. M. W. Groenen et al., “Spina bifida and genetic factors related to myo-inositol, glucose, and zinc,” Molecular Genetics and Metabolism, vol. 82, no. 2, pp. 154–161, 2004/06/01/ 2004, doi: 10.1016/j.ymgme.2004.03.007.

[30] H. Hu et al., “Correlation between congenital heart defects and maternal copper and zinc concentrations,” (in eng), Birth Defects Res A Clin Mol Teratol, vol. 100, no. 12, pp. 965–72, Dec 2014, doi: 10.1002/bdra.23284.

[31] S. F. Centofanti et al., “Maternal nutrient intake and fetal gastroschisis: A case-control study,” (in eng), Am J Med Genet A, vol. 179, no. 8, pp. 1535–1542, Aug 2019, doi: 10.1002/ajmg.a.61265.

[32] M. K. Georgieff, C. D. Petry, J. D. Wobken, and C. E. Oyer, “Liver and brain iron deficiency in newborn infants with bilateral renal agenesis (Potter’s syndrome),” (in eng), Pediatr Pathol Lab Med, vol. 16, no. 3, pp. 509–19, May-Jun 1996, doi: 10.1080/15513819609168687.

[33] D. Zeyrek, M. Soran, A. Cakmak, A. Kocyigit, and A. Iscan, “Serum copper and zinc levels in mothers and cord blood of their newborn infants with neural tube defects: a case-control study,” (in eng), Indian Pediatr, vol. 46, no. 8, pp. 675–80, Aug 2009.

[34] K. J. Rothman, L. L. Moore, M. R. Singer, U.-S. D. T. Nguyen, S. Mannino, and A. Milunsky, “Teratogenicity of High Vitamin A Intake,” New England Journal of Medicine, vol. 333, no. 21, pp. 1369–1373, 1995/11/23 1995, doi: 10.1056/NEJM199511233332101.

[35] T. W. Sadler, “The embryologic origin of ventral body wall defects,” Seminars in Pediatric Surgery, vol. 19, no. 3, pp. 209–214, 2010/08/01/ 2010, doi: 10.1053/j.sempedsurg.2010.03.006.

[36] N. M. Gude, C. T. Roberts, B. Kalionis, and R. G. King, “Growth and function of the normal human placenta,” Thrombosis Research, vol. 114, no. 5, pp. 397–407, 2004/01/01/ 2004, doi: 10.1016/j.thromres.2004.06.038.

[37] C. M. J. Tan and A. J. Lewandowski, “The Transitional Heart: From Early Embryonic and Fetal Development to Neonatal Life,” Fetal Diagnosis and Therapy, vol. 47, no. 5, pp. 373–386, 2020, doi: 10.1159/000501906.

[38] M. White, J. Arif-Pardy, and K. L. Connor, “Identification of novel nutrient-sensitive gene regulatory networks in amniocytes from fetuses with spina bifida,” (in eng), Reprod Toxicol, vol. 116, p. 108333, Dec 27 2022, doi: 10.1016/j.reprotox.2022.12.010.

[39] L. Fan, L. Xu, Y. Wang, M. Tang, and L. Liu, “Genome- and Transcriptome-Wide Characterization of bZIP Gene Family Identifies Potential Members Involved in Abiotic Stress Response and Anthocyanin Biosynthesis in Radish (Raphanus sativus L.),” (in eng), International journal of molecular sciences, vol. 20, no. 24, p. 6334, 2019, doi: 10.3390/ijms20246334.

[40] F. Xie et al., “De novo sequencing and a comprehensive analysis of purple sweet potato (Impomoea batatas L.) transcriptome,” Planta, vol. 236, no. 1, pp. 101–113, 2012/07/01 2012, doi: 10.1007/s00425-012-1591-4.

[41] L. Ma et al., “An RNA-seq-based gene expression profiling of radiation-induced tumorigenic mammary epithelial cells,” (in eng), Genomics, proteomics & bioinformatics, vol. 10, no. 6, pp. 326–335, 2012, doi: 10.1016/j.gpb.2012.11.001.

[42] Y. Wang, X. Zeng, N. J. Iyer, D. W. Bryant, T. C. Mockler, and R. Mahalingam, “Exploring the switchgrass transcriptome using second-generation sequencing technology,” (in eng), PloS one, vol. 7, no. 3, pp. e34225–e34225, 2012, doi: 10.1371/journal.pone.0034225.

[43] J.-J. Liu, R. N. Sturrock, and R. Benton, “Transcriptome analysis of Pinus monticola primary needles by RNA-seq provides novel insight into host resistance to Cronartium ribicola,” (in eng), BMC genomics, vol. 14, pp. 884–884, 2013, doi: 10.1186/1471-2164-14-884.

[44] M. White, J. Arif-Pardy, T. Van Mieghem, and K. L. Connor, “Fetal spina bifida associates with dysregulation in nutrient-sensitive placental gene networks: Findings from a matched case-control study,” (in eng), Clin Transl Sci, vol. 17, no. 1, p. e13710, Jan 2024, doi: 10.1111/cts.13710.

[45] M. P. Scott-Boyer, S. Lacroix, M. Scotti, M. J. Morine, J. Kaput, and C. Priami, “A network analysis of cofactor-protein interactions for analyzing associations between human nutrition and diseases,” Scientific Reports, vol. 6, no. 1, p. 19633, 2016/01/18 2016, doi: 10.1038/srep19633.

[46] J. D. Fischer, G. L. Holliday, and J. M. Thornton, “The CoFactor database: organic cofactors in enzyme catalysis,” (in eng), Bioinformatics, vol. 26, no. 19, pp. 2496–7, Oct 1 2010, doi: 10.1093/bioinformatics/btq442.

[47] C. The UniProt, “Update on activities at the Universal Protein Resource (UniProt) in 2013,” Nucleic Acids Research, vol. 41, no. D1, pp. D43–D47, 2013, doi: 10.1093/nar/gks1068.

[48] A. Bairoch, “The ENZYME database in 2000,” Nucleic Acids Research, vol. 28, no. 1, pp. 304–305, 2000, doi: 10.1093/nar/28.1.304.

[49] C. Andreini, I. Bertini, G. Cavallaro, G. L. Holliday, and J. M. Thornton, “Metal-MACiE: a database of metals involved in biological catalysis,” Bioinformatics, vol. 25, no. 16, pp. 2088–2089, 2009, doi: 10.1093/bioinformatics/btp256.

[50] H. Mi et al., “PANTHER version 16: a revised family classification, tree-based classification tool, enhancer regions and extensive API,” Nucleic Acids Research, vol. 49, no. D1, pp. D394–D403, 2021, doi: 10.1093/nar/gkaa1106.

[51] C. Sticht, C. De La Torre, A. Parveen, and N. Gretz, “miRWalk: An online resource for prediction of microRNA binding sites,” PLOS ONE, vol. 13, no. 10, p. e0206239, 2018, doi: 10.1371/journal.pone.0206239.

[52] R. Janky et al., “iRegulon: from a gene list to a gene regulatory network using large motif and track collections,” (in eng), PLoS Comput Biol, vol. 10, no. 7, p. e1003731, Jul 2014, doi: 10.1371/journal.pcbi.1003731.

[53] S. Parolo, S. Lacroix, J. Kaput, and M.-P. Scott-Boyer, “Ancestors’ dietary patterns and environments could drive positive selection in genes involved in micronutrient metabolism—the case of cofactor transporters,” Genes & Nutrition, vol. 12, no. 1, p. 28, 2017/10/04 2017, doi: 10.1186/s12263-017-0579-x.

[54] CDC. “Data and Statistics on Birth Defects.” https://www.cdc.gov/birth-defects/data-research/facts-stats/index.html (accessed.

[55] D. Devakumar et al., “Infectious causes of microcephaly: epidemiology, pathogenesis, diagnosis, and management,” (in eng), Lancet Infect Dis, vol. 18, no. 1, pp. e1–e13, 01 2018, doi: 10.1016/s1473-3099(17)30398-5.

[56] A. J. Barkovich, R. I. Kuzniecky, G. D. Jackson, R. Guerrini, and W. B. Dobyns, “A developmental and genetic classification for malformations of cortical development,” Neurology, vol. 65, no. 12, p. 1873, 2005, doi: 10.1212/01.wnl.0000183747.05269.2d.

[57] S. Passemard, A. M. Kaindl, and A. Verloes, “Chapter 13 - Microcephaly,” in Handbook of Clinical Neurology, vol. 111, O. Dulac, M. Lassonde, and H. B. Sarnat Eds.: Elsevier, 2013, pp. 129–141.

[58] J. B. Thornton, S. Nimer, and P. S. Howard, “The incidence, classification, etiology, and embryology of oral clefts,” (in eng), Semin Orthod, vol. 2, no. 3, pp. 162–8, Sep 1996, doi: 10.1016/s1073-8746(96)80011-9.

[59] F. A. Khan, A. Hashmi, and S. Islam, “Insights into embryology and development of omphalocele,” Seminars in Pediatric Surgery, vol. 28, no. 2, pp. 80–83, 2019/04/01/ 2019, doi: 10.1053/j.sempedsurg.2019.04.003.

[60] S. Beaudoin, “Insights into the etiology and embryology of gastroschisis,” Seminars in Pediatric Surgery, vol. 27, no. 5, pp. 283–288, 2018/10/01/ 2018, doi: 10.1053/j.sempedsurg.2018.08.005.

[61] L. S. Baskin and M. B. Ebbers, “Hypospadias: anatomy, etiology, and technique,” Journal of Pediatric Surgery, vol. 41, no. 3, pp. 463–472, 2006/03/01/ 2006, doi: 10.1016/j.jpedsurg.2005.11.059.

[62] M. M. Al-Qattan, Y. Yang, and S. H. Kozin, “Embryology of the Upper Limb,” The Journal of Hand Surgery, vol. 34, no. 7, pp. 1340–1350, 2009/09/01/ 2009, doi: 10.1016/j.jhsa.2009.06.013.

[63] D. R. Hootnick and E. M. Levinsohn, “Embryology of the lower limb demonstrates that congenital absent fibula is a radiologic misnomer,” (in eng), Anat Rec (Hoboken), vol. 305, no. 1, pp. 8–17, 01 2022, doi: 10.1002/ar.24628.

[64] Z. Miedzybrodzka, “Congenital talipes equinovarus (clubfoot): a disorder of the foot but not the hand,” Journal of Anatomy, 10.1046/j.1469-7580.2003.00147.x vol. 202, no. 1, pp. 37–42, 2003/01/01 2003, doi: 10.1046/j.1469-7580.2003.00147.x.

[65] C. M. J. Tan and A. J. Lewandowski, “The Transitional Heart: From Early Embryonic and Fetal Development to Neonatal Life,” (in eng), Fetal Diagn Ther, vol. 47, no. 5, pp. 373–386, 2020, doi: 10.1159/000501906.

[66] A. S. Ioannides and A. J. Copp, “Embryology of oesophageal atresia,” Seminars in Pediatric Surgery, vol. 18, no. 1, pp. 2–11, 2009/02/01/ 2009, doi: 10.1053/j.sempedsurg.2008.10.002.

[67] A. Kostouros, I. Koliarakis, K. Natsis, D. A. Spandidos, A. Tsatsakis, and J. Tsiaoussis, “Large intestine embryogenesis: Molecular pathways and related disorders (Review),” (in eng), Int J Mol Med, vol. 46, no. 1, pp. 27–57, Jul 2020, doi: 10.3892/ijmm.2020.4583.

[68] R. Yalavarthy and C. Parikh, “Congenital Renal Agenesis: A Review,” Saudi Journal of Kidney Diseases and Transplantation, vol. 14, no. 3, pp. 336–341, 2003.

[69] D. Kluth, “Embryology of anorectal malformations,” Seminars in Pediatric Surgery, vol. 19, no. 3, pp. 201–208, 2010/08/01/ 2010, doi: 10.1053/j.sempedsurg.2010.03.005.

[70] C. Alves and A. Rapp, “Spontaneous Abortion,” ed: StatPearls Publishing, Treasure Island (FL), 2022.

[71] H. Odland Karlsen, S. L. Johnsen, S. Rasmussen, G. Trae, H. M. T. Reistad, and T. Kiserud, “The human yolk sac size reflects involvement in embryonic and fetal growth regulation,” (in eng), Acta Obstet Gynecol Scand, vol. 98, no. 2, pp. 176–182, Feb 2019, doi: 10.1111/aogs.13466.

[72] A. Davis, T. Wiegers, R. Johnson, D. Sciaky, J. Wiegers, and C. Mattingly, “Comparative Toxicogenomics Database (CTD),” Nucleic Acids Res, 2022.

[73] M. Ringwald et al., “Mouse Genome Informatics (MGI): latest news from MGD and GXD,” (in eng), Mamm Genome, vol. 33, no. 1, pp. 4–18, 03 2022, doi: 10.1007/s00335-021-09921-0.

[74] A. Buniello et al., “The NHGRI-EBI GWAS Catalog of published genome-wide association studies, targeted arrays and summary statistics 2019,” (in eng), Nucleic acids research, vol. 47, no. D1, pp. D1005–D1012, 2019/01// 2019, doi: 10.1093/nar/gky1120.

[75] A. M. Moon, “Mouse models for investigating the developmental basis of human birth defects,” (in eng), Pediatr Res, vol. 59, no. 6, pp. 749–55, Jun 2006, doi: 10.1203/01.pdr.0000218420.00525.98.

[76] L. M. Martinelli, A. Carucci, V. J. H. Payano, K. L. Connor, and E. Bloise, “Translational Comparison of the Human and Mouse Yolk Sac Development and Function,” (in eng), Reprod Sci, vol. 30, no. 1, pp. 41–53, Jan 2023, doi: 10.1007/s43032-022-00872-8.

[77] M. Safran et al., “GeneCards Version 3: the human gene integrator,” (in eng), Database (Oxford), vol. 2010, p. baq020, Aug 05 2010, doi: 10.1093/database/baq020.

[78] G. Ge, C. A. Fernández, M. A. Moses, and D. S. Greenspan, “Bone morphogenetic protein 1 processes prolactin to a 17-kDa antiangiogenic factor,” Proceedings of the National Academy of Sciences, vol. 104, no. 24, pp. 10010–10015, 2007/06/12 2007, doi: 10.1073/pnas.0704179104.

[79] P. Kaczynski, M. P. Kowalewski, and A. Waclawik, “Prostaglandin F2α promotes angiogenesis and embryo-maternal interactions during implantation,” (in eng), Reproduction, vol. 151, no. 5, pp. 539–52, May 2016, doi: 10.1530/rep-15-0496.

[80] F. Jiang and G. E. Herman, “Analysis of Nsdhl-deficient embryos reveals a role for Hedgehog signaling in early placental development,” (in eng), Hum Mol Genet, vol. 15, no. 22, pp. 3293–305, Nov 15 2006, doi: 10.1093/hmg/ddl405.

[81] V. Ribes et al., “Rescue of cytochrome P450 oxidoreductase (Por) mouse mutants reveals functions in vasculogenesis, brain and limb patterning linked to retinoic acid homeostasis,” (in eng), Dev Biol, vol. 303, no. 1, pp. 66–81, Mar 1 2007, doi: 10.1016/j.ydbio.2006.10.032.

[82] K. S. Kottawatta, K. H. So, S. P. Kodithuwakku, E. H. Ng, W. S. Yeung, and K. F. Lee, “MicroRNA-212 Regulates the Expression of Olfactomedin 1 and C-Terminal Binding Protein 1 in Human Endometrial Epithelial Cells to Enhance Spheroid Attachment In Vitro,” (in eng), Biol Reprod, vol. 93, no. 5, p. 109, Nov 2015, doi: 10.1095/biolreprod.115.131334.

[83] L. Dhaenens et al., “Endometrial stromal cell proteome mapping in repeated implantation failure and recurrent pregnancy loss cases and fertile women,” (in eng), Reprod Biomed Online, vol. 38, no. 3, pp. 442–454, Mar 2019, doi: 10.1016/j.rbmo.2018.11.022.

[84] T. C. Genaro-Mattos, A. Anderson, L. B. Allen, Z. Korade, and K. Mirnics, “Cholesterol Biosynthesis and Uptake in Developing Neurons,” (in eng), ACS Chem Neurosci, vol. 10, no. 8, pp. 3671–3681, Aug 21 2019, doi: 10.1021/acschemneuro.9b00248.

[85] V. Nadeau, S. Guillemette, L. F. Bélanger, O. Jacob, S. Roy, and J. Charron, “Map2k1 and Map2k2 genes contribute to the normal development of syncytiotrophoblasts during placentation,” (in eng), Development, vol. 136, no. 8, pp. 1363–74, Apr 2009, doi: 10.1242/dev.031872.

[86] T. Balligand et al., “Knock-in of murine Calr del52 induces essential thrombocythemia with slow-rising dominance in mice and reveals key role of Calr exon 9 in cardiac development,” (in eng), Leukemia, vol. 34, no. 2, pp. 510–521, Feb 2020, doi: 10.1038/s41375-019-0538-1.

[87] E. Arikawa-Hirasawa, H. Watanabe, H. Takami, J. R. Hassell, and Y. Yamada, “Perlecan is essential for cartilage and cephalic development,” (in eng), Nat Genet, vol. 23, no. 3, pp. 354–8, Nov 1999, doi: 10.1038/15537.

[88] B. Chandrasekharan, C. Montllor-Albalate, A. E. Colin, J. L. Andersen, Y. C. Jang, and A. R. Reddi, “Cu/Zn Superoxide Dismutase (Sod1) regulates the canonical Wnt signaling pathway,” (in eng), Biochem Biophys Res Commun, vol. 534, pp. 720–726, Jan 1 2021, doi: 10.1016/j.bbrc.2020.11.011.

[89] T. L. Capasso et al., “BMP10-mediated ALK1 signaling is continuously required for vascular development and maintenance,” (in eng), Angiogenesis, vol. 23, no. 2, pp. 203–220, May 2020, doi: 10.1007/s10456-019-09701-0.

[90] Y. Chen et al., “Type-I collagen produced by distinct fibroblast lineages reveals specific function during embryogenesis and Osteogenesis Imperfecta,” (in eng), Nat Commun, vol. 12, no. 1, p. 7199, Dec 10 2021, doi: 10.1038/s41467-021-27563-3.

[91] A. Hafez, R. Squires, A. Pedracini, A. Joshi, R. E. Seegmiller, and J. T. Oxford, “Col11a1 Regulates Bone Microarchitecture during Embryonic Development,” (in eng), J Dev Biol, vol. 3, no. 4, pp. 158–176, 2015, doi: 10.3390/jdb3040158.

[92] J. Cui, K. S. O’Shea, A. Purkayastha, T. L. Saunders, and D. Ginsburg, “Fatal haemorrhage and incomplete block to embryogenesis in mice lacking coagulation factor V,” (in eng), Nature, vol. 384, no. 6604, pp. 66–8, Nov 7 1996, doi: 10.1038/384066a0.

[93] X. Rong et al., “Glutathione peroxidase 4 inhibits Wnt/β-catenin signaling and regulates dorsal organizer formation in zebrafish embryos,” (in eng), Development, vol. 144, no. 9, pp. 1687–1697, May 1 2017, doi: 10.1242/dev.144261.

[94] M. Sun et al., “Collagen V is a dominant regulator of collagen fibrillogenesis: dysfunctional regulation of structure and function in a corneal-stroma-specific Col5a1-null mouse model,” (in eng), J Cell Sci, vol. 124, no. Pt 23, pp. 4096–105, Dec 1 2011, doi: 10.1242/jcs.091363.

[95] D. Zhou et al., “hiPSC Modeling of Lineage-Specific Smooth Muscle Cell Defects Caused by TGFBR1(A230T) Variant, and Its Therapeutic Implications for Loeys-Dietz Syndrome,” (in eng), Circulation, vol. 144, no. 14, pp. 1145–1159, Oct 5 2021, doi: 10.1161/circulationaha.121.054744.

[96] J. Liu et al., “Talin1 regulates integrin turnover to promote embryonic epithelial morphogenesis,” (in eng), Mol Cell Biol, vol. 31, no. 16, pp. 3366–77, Aug 2011, doi: 10.1128/mcb.01403-10.

[97] K. Long, L. Moss, L. Laursen, L. Boulter, and C. Ffrench-Constant, “Integrin signalling regulates the expansion of neuroepithelial progenitors and neurogenesis via Wnt7a and Decorin,” (in eng), Nat Commun, vol. 7, p. 10354, Feb 3 2016, doi: 10.1038/ncomms10354.

[98] X. Yu et al., “The Cytokine TGF-β Promotes the Development and Homeostasis of Alveolar Macrophages,” (in eng), Immunity, vol. 47, no. 5, pp. 903–912.e4, Nov 21 2017, doi: 10.1016/j.immuni.2017.10.007.

[99] L. D. Derycke and M. E. Bracke, “N-cadherin in the spotlight of cell-cell adhesion, differentiation, embryogenesis, invasion and signalling,” (in eng), Int J Dev Biol, vol. 48, no. 5-6, pp. 463–76, 2004, doi: 10.1387/ijdb.041793ld.

[100] Y. Komiya and R. Habas, “Wnt signal transduction pathways,” (in eng), Organogenesis, vol. 4, no. 2, pp. 68–75, Apr 2008, doi: 10.4161/org.4.2.5851.

[101] L. M. R. Ferreira, A. M. Li, T. L. Serafim, M. C. Sobral, M. C. Alpoim, and A. M. Urbano, “Intermediary metabolism: An intricate network at the crossroads of cell fate and function,” (in eng), Biochim Biophys Acta Mol Basis Dis, vol. 1866, no. 10, p. 165887, Oct 01 2020, doi: 10.1016/j.bbadis.2020.165887.

[102] A. Casanova, A. Wevers, S. Navarro-Ledesma, and L. Pruimboom, “Mitochondria: It is all about energy,” (in eng), Front Physiol, vol. 14, p. 1114231, 2023, doi: 10.3389/fphys.2023.1114231.

[103] S. Wang et al., “Transferrin receptor 1-mediated iron uptake plays an essential role in hematopoiesis,” (in eng), Haematologica, vol. 105, no. 8, pp. 2071–2082, Aug 2020, doi: 10.3324/haematol.2019.224899.

[104] R. J. Cousins, “A role of zinc in the regulation of gene expression,” (in eng), Proc Nutr Soc, vol. 57, no. 2, pp. 307–11, May 1998, doi: 10.1079/pns19980045.

[105] R. S. MacDonald, “The role of zinc in growth and cell proliferation,” (in eng), J Nutr, vol. 130, no. 5S Suppl, pp. 1500S–8S, 05 2000, doi: 10.1093/jn/130.5.1500S.

[106] M. Figiel, A. K. Górka, and A. Górecki, “Zinc Ions Modulate YY1 Activity: Relevance in Carcinogenesis,” (in eng), Cancers (Basel), vol. 15, no. 17, Aug 30 2023, doi: 10.3390/cancers15174338.

[107] G. U. Martinez-Ruiz, A. Morales-Sanchez, and A. F. Pacheco-Hernandez, “Roles Played by YY1 in Embryonic, Adult and Cancer Stem Cells,” (in eng), Stem Cell Rev Rep, vol. 17, no. 5, pp. 1590–1606, Oct 2021, doi: 10.1007/s12015-021-10151-9.

[108] H. Vannucchi and F. S. Moreno, “Interaction of niacin and zinc metabolism in patients with alcoholic pellagra,” (in eng), Am J Clin Nutr, vol. 50, no. 2, pp. 364–9, Aug 1989, doi: 10.1093/ajcn/50.2.364.

[109] H. Vannucchi, M. D. Kutnink, M. Sauberlich, and E. Howerde, “Interaction among niacin, vitamin B6 and zinc in rats receiving ethanol,” (in eng), Int J Vitam Nutr Res, vol. 56, no. 4, pp. 355–62, 1986.

[110] C. T. Mai et al., “National population-based estimates for major birth defects, 2010-2014,” (in eng), Birth Defects Res, vol. 111, no. 18, pp. 1420–1435, Nov 01 2019, doi: 10.1002/bdr2.1589.

[111] M. El-Chouli et al., “Time Trends in Simple Congenital Heart Disease Over 39 Years: A Danish Nationwide Study,” (in eng), J Am Heart Assoc, vol. 10, no. 14, p. e020375, Jul 20 2021, doi: 10.1161/JAHA.120.020375.

