## Supplementary Figures for "Identification of novel nutrient sensitive human yolk sac functions required for embryogenesis"

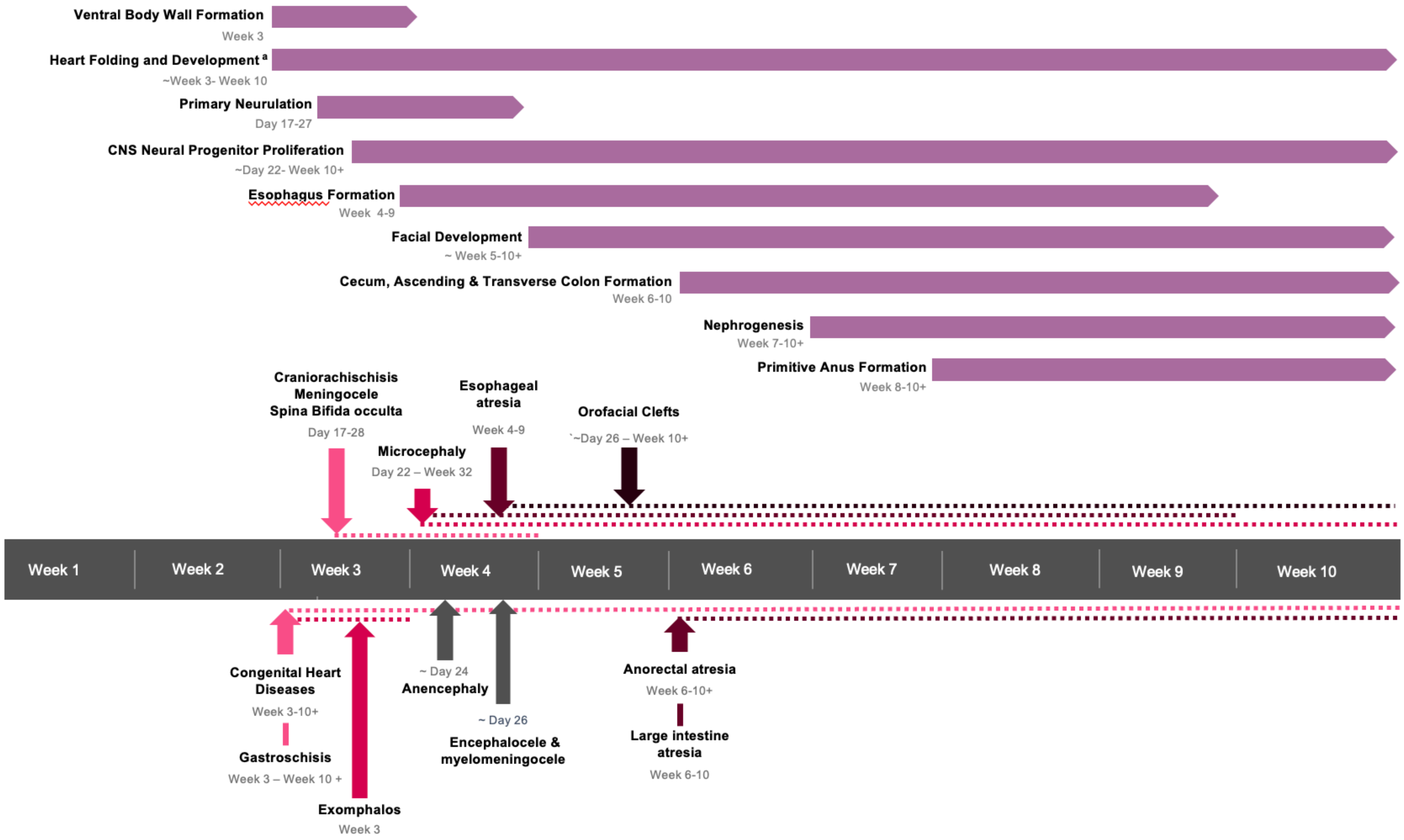


**Supplementary Figure S1. Developmental stages and potential etiological time periods for major congenital diseases.** [a] By the end of the 5th week of gestation, heart folding completes, and by the end of the 7th week, the septum intermedium, clear atria, and auricles are formed, and the ventricular septum stops growing. By the 10th week, the aortic and pulmonary ventricles and outflow tracts are entirely separated, the coronary sinus is formed, and the semilunar valves are complete.

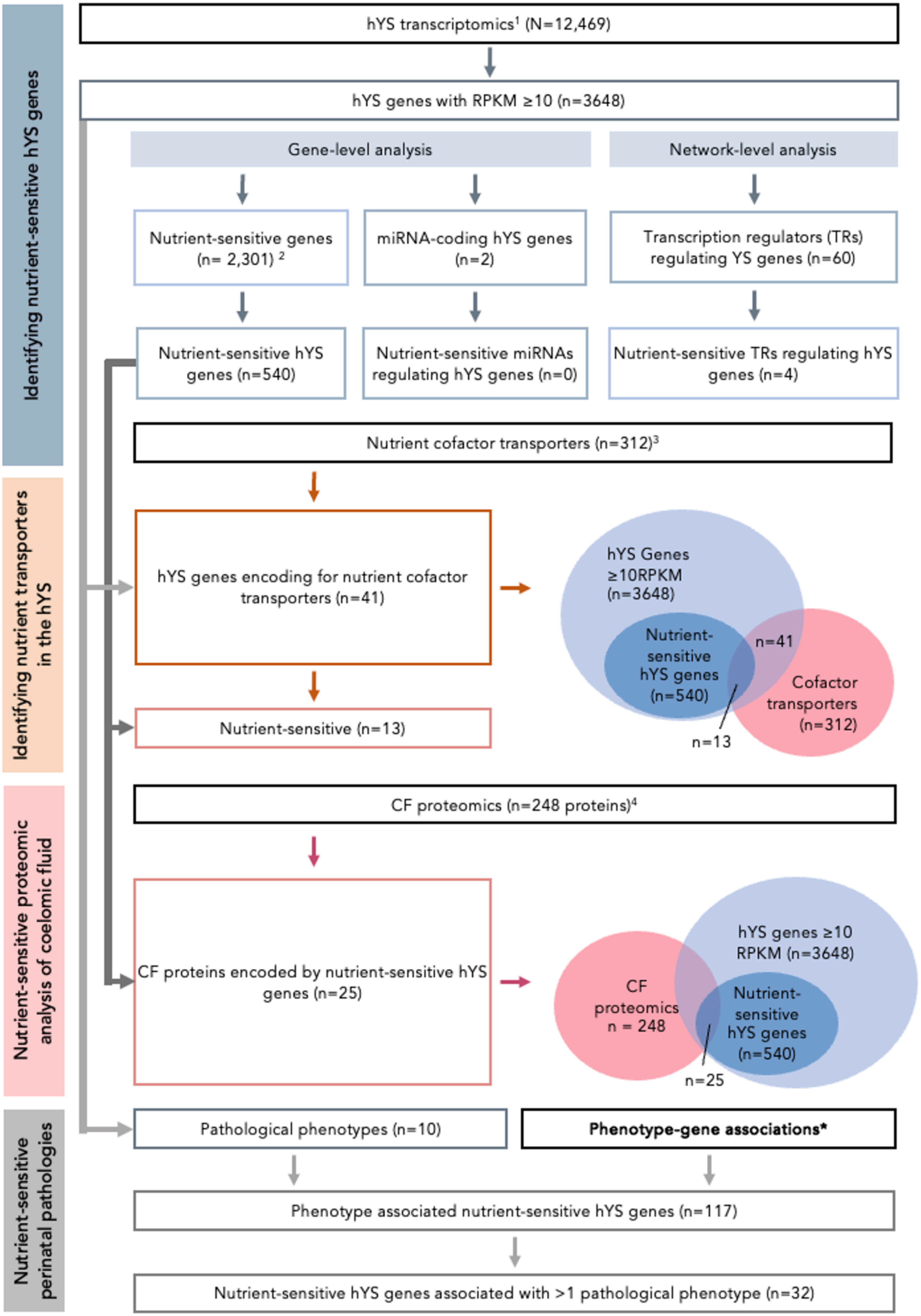


**Supplementary Figure S2. Flow diagram indicating levels of human yolk sac (hYS) gene-nutrient transcriptomic-, coelomic fluid (CF) proteomic-, and congenital disease analysis**. YS = yolk sac; CF = coelomic fluid; NTD = neural tube defect; CHD = congenital heart defect; RPKM = reads per kilobase of transcript, per million mapped reads. [1]Cindrova-Davis, 2017. [2]Scott-Boyer, 2016. [3]Parolo, 2017. [4]Aiello, 2018. *See hYS pathological phenotype analysis methods section for details on associations origin.

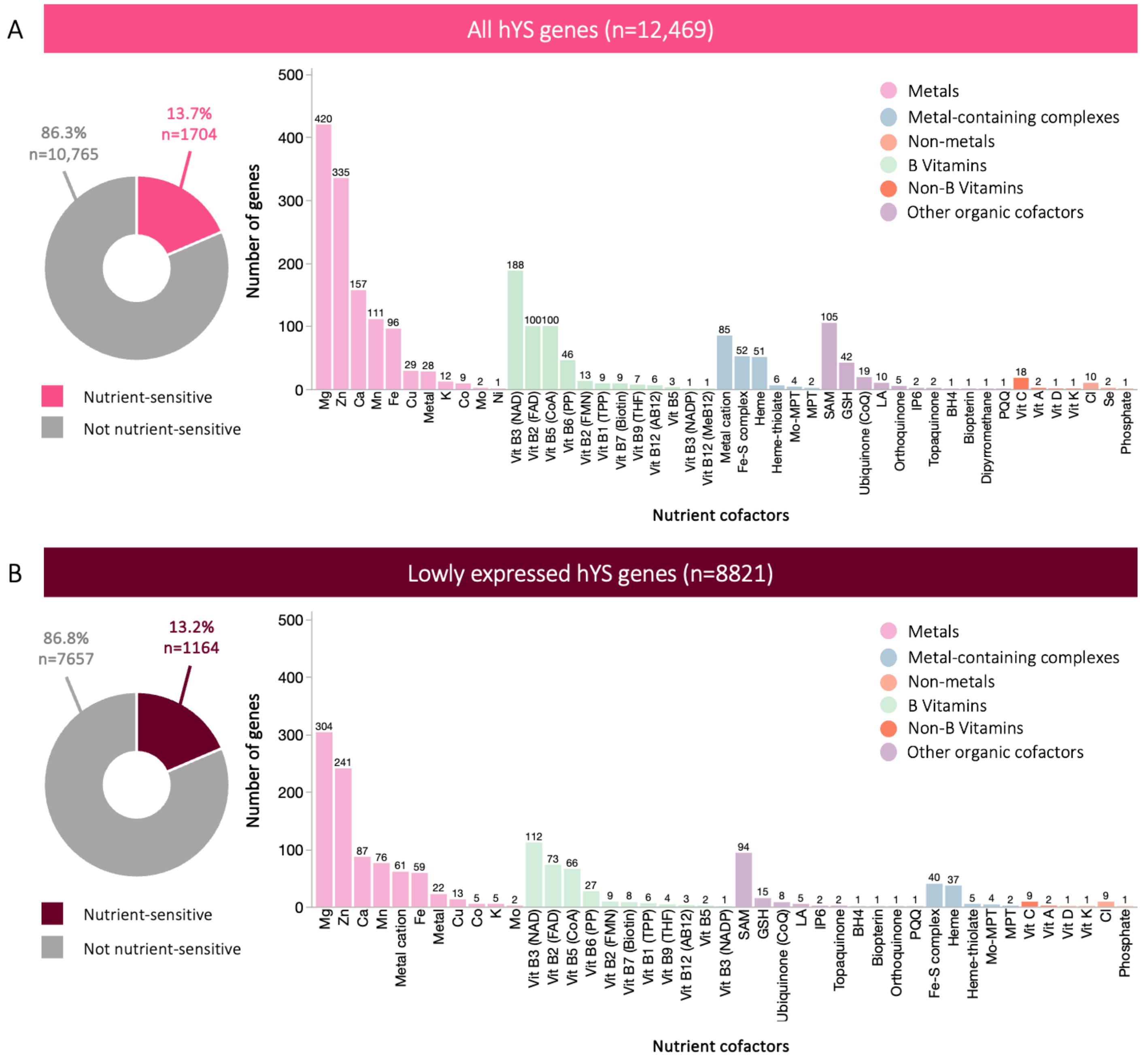


**Supplementary Figure S3. Nutrient-sensitive genes in hYS genes stratified by expression group.** Proportion of hYS genes that are nutrient-sensitive (left column; donut charts) to the nutrients indicated in the histograms (right column; bar charts), for (A) all hYS genes, and (B) lowly expressed hYS genes. Numbers above the histogram bars = total number of genes sensitive to that nutrient. RPKM = Reads per kilobase of transcript, per million mapped reads; SAM = S-Adenosyl methionine; GSH = glutathione; Mo = molybdenum; MPT = molybdopterins.

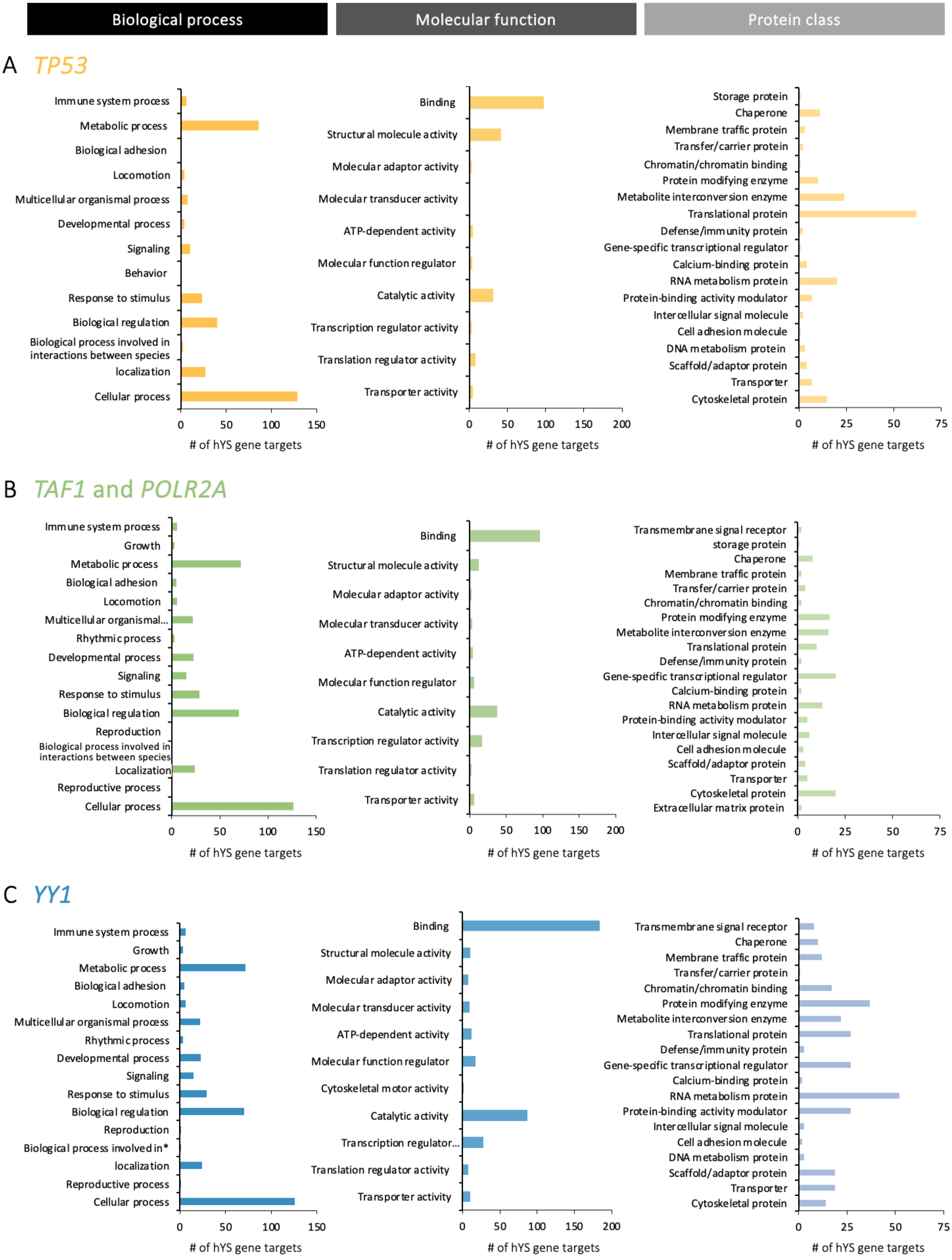


**Supplementary Figure S4. Functional analysis of highly expressed hYS genes regulated by nutrient-sensitive transcriptional regulators.** Biological processes, molecular functions, and protein classes of hYS genes predicted to be regulated by (A) *TP53* (zinc-sensitive), (B) *TAF1* and *POLR2A* (zinc- and magnesium-sensitive), and (C) *YY1* (zinc-sensitive).
