## Supplementary Table S3 for "Identification of novel nutrient sensitive human yolk sac functions required for embryogenesis"

**Supplementary Table S3.** Genes expressed in the human yolk sac that encode a micronutrient transporter.

| Gene Name | Gene Symbol | Transported Nutrient | Nutrient Cofactor | Known Target/Function |
|---|---|---|---|---|
| ATPase sarcoplasmic/endoplasmic reticulum Ca2+ transporting 2 | ATP2A2 | Calcium | Calcium, Magnesium | Encodes an ATPase which transports calcium from the cytosol to the sarcoplasmic reticulum lumen. Involved in autophagy, and regulation of the contraction/relaxation cycle in muscle cells. |
| ATPase plasma membrane Ca2+ transporting 1 | ATP2B1 | Calcium | Magnesium | Encodes an ATPase which transports calcium from the cytoplasm to the extracellular area. Involved in blood pressure regulation, bone mineralization, osteoclast differentiation, and synaptic transmission. |
| ATPase Plasma Membrane Ca2+ Transporting 4 | ATP2B4 | Calcium | Magnesium | Encodes an ATPase that catalyzes ATP hydrolysis, facilitating the transport of calcium out of the cell. Regulates sperm cell calcium homeostasis. |
| Solute carrier family 31 member 1 | SLC31A1 | Copper | - | Encodes a high affinity copper transporter found in the cell membrane and is involved in the uptake of dietary copper intake. |
| Cytochrome B reductase 1 | CYBRD1 | Iron | Heme | Encodes a cytochrome B reductase protein involved in transferring an electron to Fe(3+), thus playing a role in physiological iron absorption. |
| Ferritin heavy chain 1 | FTH1 | Iron | Iron | Encodes the heavy subunit of ferritin, which is a protein that stores iron in a soluble, non-toxic, available form. Involved in regulation of iron uptake and release into various tissues. |
| Ferritin light chain | FTL | Iron | Iron | Encodes the light chain of ferritin, which is a protein that stores iron in a soluble, non-toxic, available form. Involved in regulation of iron uptake and release into various tissues. |
| Hephaestin | HEPH | Iron | Copper | Encodes a ferroxidase protein, which oxidizes Fe(2+). Involved in the transport of iron from epithelial cells to the lumen of the circulatory system. |
| Heme oxygenase 1 | HMOX1 | Iron | Heme, Vitamin B3 | Encodes a heme oxygenase, which cleaves heme to form biliverdin (later converted to bilirubin), during which it releases the hemes iron as a ferrous ion. |
| Hemopexin | HPX | Iron | - | Encodes a plasma glycoprotein, which binds to heme and transports it from the plasma to the liver for breakdown and iron recycling. |
| Solute carrier family 25 member 28 | SLC25A28 | Iron | - | Encodes a mitochondrial iron transporter that mediates uptake of iron into the mitochondrion. |

| | | | | |
|---|---|---|---|---|
| Solute carrier family 25 member 37 | SLC25A37 | Iron | - | Encodes a mitochondrial iron transporter that specifically mediates the uptake of iron into the developing erythrocytes and is thus important in heme formation. |
| Solute carrier family 39 member 14 | SLC39A14 | Iron | - | Encodes a protein involved in the transfer of nontransferrin-bound iron and is embedded within the cell membrane. Also involved in magnesium and zinc transfer |
| Solute carrier family 40 member 1 | SLC40A1 | Iron | - | Encodes a cell membrane transporter responsible for the export of iron form duodenal epithelial cells. Plays a role in transferring iron from intestinal, splenic, and hepatic cells into the blood. Also involved in iron recycling in macrophages. |
| Transferrin | TF | Iron | Iron | Encodes a transferrin protein, which is involved in iron binding and the transport of iron from sites of absorption and heme degradation to sites of iron utilization. |
| Transferrin receptor | TFRC | Iron | - | Encodes a transferrin receptor, which binds to transferrin and mediates the transport of iron into and out of cells via receptor-mediated endocytosis. |
| Magnesium transporter 1 | MAGT1 | Magnesium | - | Encodes a ubiquitously expressed magnesium transporter located in the cell membrane. |
| NIPA magnesium transporter 2 | NIPA2 | Magnesium | - | Encodes a selective magnesium (2+) transporter. |
| Solute carrier family 39 member 7 | SLC39A7 | Manganese | - | Encodes a protein which transports zinc (2+) from the golgi apparatus or endoplasmic reticulum to the cytosol. Also has manganese transporter activity. |
| Selenoprotein P | SEPP1 | Selenium | - | Encodes a selenoprotein highly expressed in the liver. Plays a role in extracellular antioxidant defence but also in the transport of selenium. |
| Solute carrier family 30 member 1 | SLC30A1 | Zinc | - | Encodes a protein which enables calcium channel inhibition and has zinc ion transmembrane activity (predicted to be involved in zinc export). |
| Solute carrier family 39 member 1 | SLC39A1 | Zinc | - | Encodes a transporter protein primarily responsible for the endogenous zinc uptake. |
| Solute carrier family 39 member 13 | SLC39A13 | Zinc | - | Encodes a transmembrane protein which acts as a zinc-influx transporter. |
| Solute carrier family 39 member 5 | SLC39A5 | Zinc | - | Encodes a zinc-influx transporter and thus plays an important role in maintaining intracellular zinc levels. Plays a role in eye development. |
| Solute carrier family 39 member 6 | SLC39A6 | Zinc | - | Encodes a zinc-influx transporter and thus plays an important role in maintaining intracellular zinc levels. |
| Solute carrier family 39 member 9 | SLC39A9 | Zinc | - | Encodes a transmembrane zinc-influx transporter. |
| FLVCR heme transporter 1 | FLVCR1 | Heme | - | Encodes a heme transporter that exports cytoplasmic heme from the mitochondrion to the cytoplasm and is essential in erythroid differentiation (protects erythrocytes from heme toxicity). |
| LMBR1 domain containing 1 | LMBRD1 | Vitamin B12 | - | Encodes a lysosomal membrane chaperone protein that exports vitamin B12 from the lysosome to the cytosol. |
| Solute carrier family 16 member 1 | SLC16A1 | Vitamin B7 | - | Encodes a proton-linked monocarboxylate transporter, transporting molecules such as lactate, pyruvate, or biotin (vitamin B7). |
| Folate receptor beta | FOLR2 | Vitamin B9 | Vitamin B6 | Encodes a folate receptor which binds to folic acid and its derivatives, mediating the transport to the cell's interior. |

| | | | | |
|---|---|---|---|---|
| Solute carrier family 19 member 2 | SLC19A2 | Vitamin B1 | - | Encodes a high-affinity thiamine transporter. |
| Retinol binding protein 1 | RBP1 | Vitamin A | Vitamin B6 | Encodes a protein which binds retinol in the cytoplasm (primarily in the liver) and then transports it to peripheral tissues. |
| Retinol binding protein 2 | RBP2 | Vitamin A | Vitamin B6 | Encodes a protein primarily found in the small intestine responsible for the uptake of retinol. |
| Retinol binding protein 4 | RBP4 | Vitamin A | - | Encodes a protein that binds to retinol in the liver and transports it to peripheral tissues. |
| Anoctamin 1 | ANO1 | Chloride | - | Encodes a calcium-activated chloride channel, which is responsible for transporting chloride across epithelial membranes and in smooth muscle contraction. |
| Chloride voltage-gated Channel 3 | CLCN3 | Chloride | Chloride | Encodes a voltage gated chloride channel, present in all cell types and in intracellular vesicles. |
| Solute carrier family 12 Member 7 | SLC12A7 | Chloride | - | Encodes a protein involved in cell volume homeostasis by mediating potassium and chloride transport in response to cell swelling. |
| Solute carrier family 4 Member 1 | SLC4A1 | Chloride | - | Encodes an anion transporter in the erythrocyte membrane responsible for maintaining erythrocyte shape. |
| Solute carrier family 4 member 2 | SLC4A2 | Chloride | - | Encodes a plasma membrane protein responsible for the transport of anions. |
| Solute carrier family 20 member 1 | SLC20A1 | Phosphate | - | Encodes a sodium-phosphate symporter that absorbs phosphate from the interstitial fluid for internal uses. |
| Solute carrier family 34 member 2 | SLC34A2 | Phosphate | - | Encodes a pH-sensitive, sodium dependent phosphate transporter, which transports phosphate inside of a cell via sodium symport transport. |

All key information on gene (protein) functions and processes were obtained from genecards.org [77].
